# Exposures and conditions prior to age 16 are associated with dementia status among adults in the United States Health and Retirement Study

**DOI:** 10.1101/2024.08.15.24312018

**Authors:** Scarlet Cockell, Herong Wang, Kelly S. Benke, Erin B. Ware, Kelly M. Bakulski

## Abstract

**Background:** Dementia susceptibility likely begins years before symptoms. Early life has not been comprehensively tested for dementia associations.

**Method:** In the US Health and Retirement Study (normal baseline cognition; n=16,509; 2008-2018 waves), 31 exposures before age 16 were retrospectively assessed with ten-year incident cognitive status (dementia, impaired, normal). Using parallel logistic models, each exposure was tested with incident cognition, adjusting for sex, baseline age, follow-up, race/ethnicity, personal/parental education.

**Result:** 14.5% had incident impairment and 5.3% had dementia. Depression was associated with 1.71 (95%CI:1.28,2.26) times higher odds of incident impairment, relative to normal cognition. Headaches/migraines were associated with 1.63 (95%CI:1.18,2.22) times higher odds of incident impairment. Learning problems were associated with 1.75 (95%CI:1.05,2.79) times higher odds of incident impairment. Childhood self-rated health of fair (1.86, 95%CI:1.27,2.64) and poor (3.39, 95%CI:1.91,5.82) were associated with higher incident dementia odds, relative to excellent.

**Conclusion:** Early life factors may be important for impairment or dementia, extending the relevant risk window.

## Introduction

Dementia, a progressive neurogenerative disease, impacts an estimated 55 million worldwide, and approximately 10 million new cases are reported yearly.^1–4^ With the number of individuals 65 and older growing within the United States, dementia incidence and annual costs attributable to dementia are anticipated to increase.^1,4^ Disease prevalence is highest among adults 65 and older, making age the greatest risk factor for Alzheimer’s and other dementias.^1^ As multidisciplinary dementia research continues, medications aimed at slowing disease progression have been developed, and identifying risk factors for prevention is a key public health approach.^5,6^

To date, 12 dementia risk factors have been identified in mid and late life which may prevent or postpone up to 40% of incident dementia cases: depression, smoking, physical inactivity, diabetes, social isolation, air pollution, hypertension, hearing impairment, obesity, excessive alcohol use, and brain injury.^6–10^ Much focus has been given to the mid and late-life periods for exposures critical to the development of Alzheimer’s disease and dementia. However, widening the scope of research into childhood and adolescence may unearth additional modifiable factors, allowing for novel prevention and intervention measures at the earliest opportunity.

Cognitive changes are present and detectable years before individuals meet the clinical criteria for dementia.^3,7–9^ Therefore, beginning stages of neurodegeneration are present in the brain long before symptoms are apparent, however exactly when these changes begin is unclear.^5,8–10^ To precede clinical, subclinical, or pathological changes investigations must consider earlier windows of susceptibility. Few strong childhood environmental risk factors have been identified for dementia, likely due to the long duration between exposure and onset, which presents challenges for measurement in the same Individuals.^8,11–13^ Due to these difficulties, investigations into how social and environmental factors in early life influence disease development are incomplete.^4–7,12^ Screening the early life period may be instrumental in identifying the earliest exposure window to influence dementia risk.

In a large and diverse sample of older adults from the United States Health and Retirement Study (HRS), this study aims to screen 31 early-life exposures measured at baseline with ten years of cognitive follow-up for their association with incident dementia or incident cognitive impairment non-dementia later in life. Using an exposure-wide association study (ExWAS) framework (PMID: 32105887, 38344436), this study investigates a diverse group of exposures in the early life stage and their association with dementia in late life to interrogate an earlier window of susceptibility.^14,15^ We hypothesize that previously identified modifiable risk factors may impact cognition in early life and thus contribute to the development of dementia. The long pre-clinical phase of dementia provides many opportunities for intervention, and through this study, we hope to elucidate additional risk factors to identify prevention periods in early life.

## Methods

### Study sample

The United States HRS includes a nationally representative sample of individuals aged 50 years and older who have been interviewed biennially since 1992 and covers topics like income, wealth, health, health services use, work, retirement, and family dynamics. The HRS uses a panel cohort study design. An overview of this study has been given in detail previously.^16,17^ The HRS is implemented by the Institute for Social Research at the University of Michigan and is sponsored by the National Institute on Aging (U01 AG009740). Individuals provided written informed consent at the time of study participation. These secondary data analyses were approved by the University of Michigan Institutional Review Board (HUM00128220). HRS data are publicly available (https://hrs.isr.umich.edu). Our analysis used self-reported exposures and conditions that occurred before 16 years old collected from the 2008 to 2014 waves (baseline measure), and cognition data from the 2008 to 2018 waves (follow-up) for the same individuals (**Supplementary Figure 1**).

### Early life exposure and condition measures

In 2008, a new HRS questionnaire inquired retrospectively about early life exposures or conditions before age 16. Each of these 31 exposures were included in this study. Exposure variables included infection medical history (infection with measles, mumps, chicken pox, allergies), neurologic factors (headache, ear problems, depression, emotional and psychological problems, learning disorder, concussion, epilepsy, drug smoking, or alcohol use), respiratory factors (parental smoking, asthma, respiratory condition), sensory deficits (speech, vision), other physical factors (heart circulatory and blood conditions, disability, stomach condition, self-reported health, digestive system, musculoskeletal and connective tissue conditions, any other conditions), and child home environment (lived in rural area, missed school, lived with grandparents, father unemployment, mother work, family financial situation, family financial help). Individuals who did not respond to the 2008 questionnaire were invited to respond at each subsequent visit through 2014. All Individuals who answered at least one of the 31 questions were included. Although these questions were intended to be asked once, some subjects (n=202) had two responses between 2008 and 2014. If the interviewee answered differently between responses, the Individual was excluded for that exposure. Due to small sample sizes, for the question “Did your mother work during childhood?” Individuals with the response “Never lived with mother/mother was not alive” (n=31) were excluded. Similarly, for the prompt “Rate your family financial situation,” Individual s with the response “It varied” (n=107) were excluded. We considered an Individual’s baseline measure to be the wave in which they completed the early life exposure questionnaire.

Eight summary categories were created to aid in the interpretation of the individual exposures. Original question wording, internal labels, and summary variable membership can be found in **supplementary table 1**. The summary exposure category *Inflammatory and infectious upper respiratory conditions* included three variables: “Allergic conditions”, “Allergic conditions disability” and “Allergic conditions other”. Likewise, the summary exposure category *Psychological conditions* included three variables: “Emotional or psychological conditions”, “Emotional or psychological conditions disability” and “Psychological or emotional conditions other”. For the summary exposure category *Neurological conditions* three variables were combined: “Epilepsy or seizures”, “Neurological and sensory conditions disability” and “Neurological and sensory conditions other.” The creation of the *Childhood substance use* summary exposure category combines two variables: “Drug or alcohol use” and “Regular cigarette smoking”. The summary exposure category *Heart conditions* is a combination of three variables: “High blood pressure”, “Heart conditions disability” and “Heart conditions other.” The *Digestive system conditions* summary exposure unites two variables: “Digestive system conditions disability” and “Digestive system conditions other”. The *Musculoskeletal and connective tissue conditions* summary category included two variables: “Musculoskeletal and connective tissue conditions disability” and “Musculoskeletal and connective tissue conditions other”. Finally, the summary exposure category *Childhood health conditions* included nine variables: “Endocrine, metabolic, and nutritional conditions disability”, “Endocrine, metabolic, and nutritional conditions other”, “Reproductive system and prostate conditions disability”, “Reproductive system and prostate conditions other”, “Cancers, tumors, skin conditions disability”, “Cancers, tumors, skin conditions other”, “Diabetes”, “Other health conditions”, and “Other disability”. All summary exposure variables were created using the same method. If an Individual was missing a response for any original variable, the summary variable was set as missing. Among complete responses, if the Individual responded as “Yes” to any original variable, the response was coded as “Yes,” and if the Individual responded “No” to all original variables, the response was coded as “No”.

### Cognitive status measures

For cognitive status measures, we used the HRS Cross-Wave Imputation of Cognitive Functioning Measures for the years 2008 to 2018. Self-respondent cognitive status was assessed on a 27-point scale by summing an immediate and delayed 10-noun free recall test (0 to 20 points), a serial 7 subtraction test (0 to 5 points), and a backward count from 20 test (0 to 2 points).^17,18^ Respondents were classified into normal cognition (score range: 12-27), cognitive impairment non-dementia (7-11), and dementia (0-6).^18^ For respondents represented by a proxy (baseline N= 153, follow-up N= 812), an alternative 11-point scale was used.^18^ The proxy’s assessment of the respondent’s memory ranged from excellent to poor (0 to 4 points), the number of instrumental activities of daily living with limitations (0 to 5 points), and the survey interviewer’s assessment of whether the respondent had difficulty completing the interview because of a cognitive limitation (0 to 2 points).^18^ Established cutoffs based on proxy scores classified the respondent into one of the three categories: normal cognition (0-2), cognitive impairment non-dementia (3-5), or dementia (6-11).^18^ status).

### Covariate measures

Self-response to demographic questions including sex (male, female), race/ethnicity category (non-Hispanic white, non-Hispanic black, Hispanic, and non-Hispanic other), own education (years), and parental highest degree of education (high school or higher, less than high school) were included. The parental education covariate was created by using the maximum of either the maternal or paternal educational attainment.^19^ Enrollment self-response to birthdate was used to calculate age at baseline (years) and the age at the most recent cognition measure (years). Follow-up time was calculated as the time (years) between the baseline exposure collection and the most recent cognition measure as a continuous variable.

### Statistical analysis

Individuals were excluded from the analytic sample for missing exposure, covariate (sex, age, race/ethnicity, educational attainment, parental education), or outcome measures. Individuals with prevalent dementia or cognitive impairment non-dementia at baseline were also excluded. Individual inclusion and exclusion were visualized using a flow chart. Due to differences in missingness between exposures, the sample size was allowed to vary by exposure. The distributions in the included (for any exposure) and excluded samples were described using mean and standard deviation for continuous variables as well as count and frequency for categorical variables. We compared the sample distributions using Kruskal-Wallis rank sum test for continuous variables and Fisher’s test for categorical variables. Due to the non-normal left-skewed distribution of continuous variables and the presence of our three-level outcome variable, the Kruskal-Wallis rank sum test was used for the comparison of sample distributions for continuous variables. Fisher’s test was used for comparison of sample distribution for categorical variables due to the small exposure sample size for some exposure variables. Within the analytic sample, we described the distributions of variables by cognitive status after follow-up, using mean and standard deviation for continuous variables as well as count and frequency for categorical variables.

To assess exposure-specific associations with cognitive status, we used an ExWAS design and performed parallel multivariable logistic regression.^14,15^ Two sets of logistic regression models were used (dementia versus normal cognition; cognitive impairment non-dementia versus normal cognition). We first presented unadjusted crude associations between each exposure and cognitive status. Then our primary models were adjusted for sex, race/ethnicity, age at baseline, education, parental education, and time from baseline to cognitive follow-up. We presented the regression results as an odds ratio (OR), 95% confidence interval (CI), and p-value. The Benjamini-Hochberg procedure was applied to account for multiple comparisons, and FDR values were calculated.^20^ To support our goal of screening a diverse selection of exposures for association with cognitive status we used two criteria (stringent, permissive). Our stringent criteria consider an FDR<0.05 statistically significant, while the permissive criteria consider a p-value<0.05 sufficient regardless of FDR. Permissive criteria were included for exploratory purposes to support our primary objective of screening a diverse set of early-life factors for association with incident cognitive status.

To assess the joint effect across exposures within a summary category, we used the metafor package for random-effect meta-analyses to estimate the pooled odds ratios from all exposures under the same category.^21^ We assessed heterogeneity within the category using I^2^. We visualized odds ratios for all individual exposures and pooled odds ratios for summary category exposures using forest plots.

### Sensitivity analyses

Following primary analyses, we assessed an additional model using an alternative impaired cognition definition. Individuals with cognitive impairment non-dementia and dementia were combined to create a binary outcome variable of normal cognition or any cognitive impairment. All 31 exposures were individually assessed for their relationship with any cognitive impairment adjusting for age, sex, race/ethnicity, personal education, parental education, and time from baseline to cognitive follow-up. Statistical significance was assessed using only our stringent criteria (FDR<0.05). All statistical analyses were conducted using R 3.5.3. Code to conduct analyses is available https://github.com/bakulskilab.

## Results

### Study sample descriptive statistics

There were 22,018 individuals eligible for this study and our included analytic sample size was 16,509 (**Figure 1**). We excluded those with cognitive impairment at baseline (n=4,320) and those without follow-up cognition visit information (n=1,189). Relative to the included sample, those excluded were older (mean 69 years), had fewer years of schooling (mean 11.1), were more likely to have had parents who completed fewer school years, and were more likely to be of a race/ethnicity other than Non-Hispanic White (Non-Hispanic Black: 25%, Hispanic: 18%, Non-Hispanic other 3.4%) (**Supplemental Table 2).**

**Figure 1.**
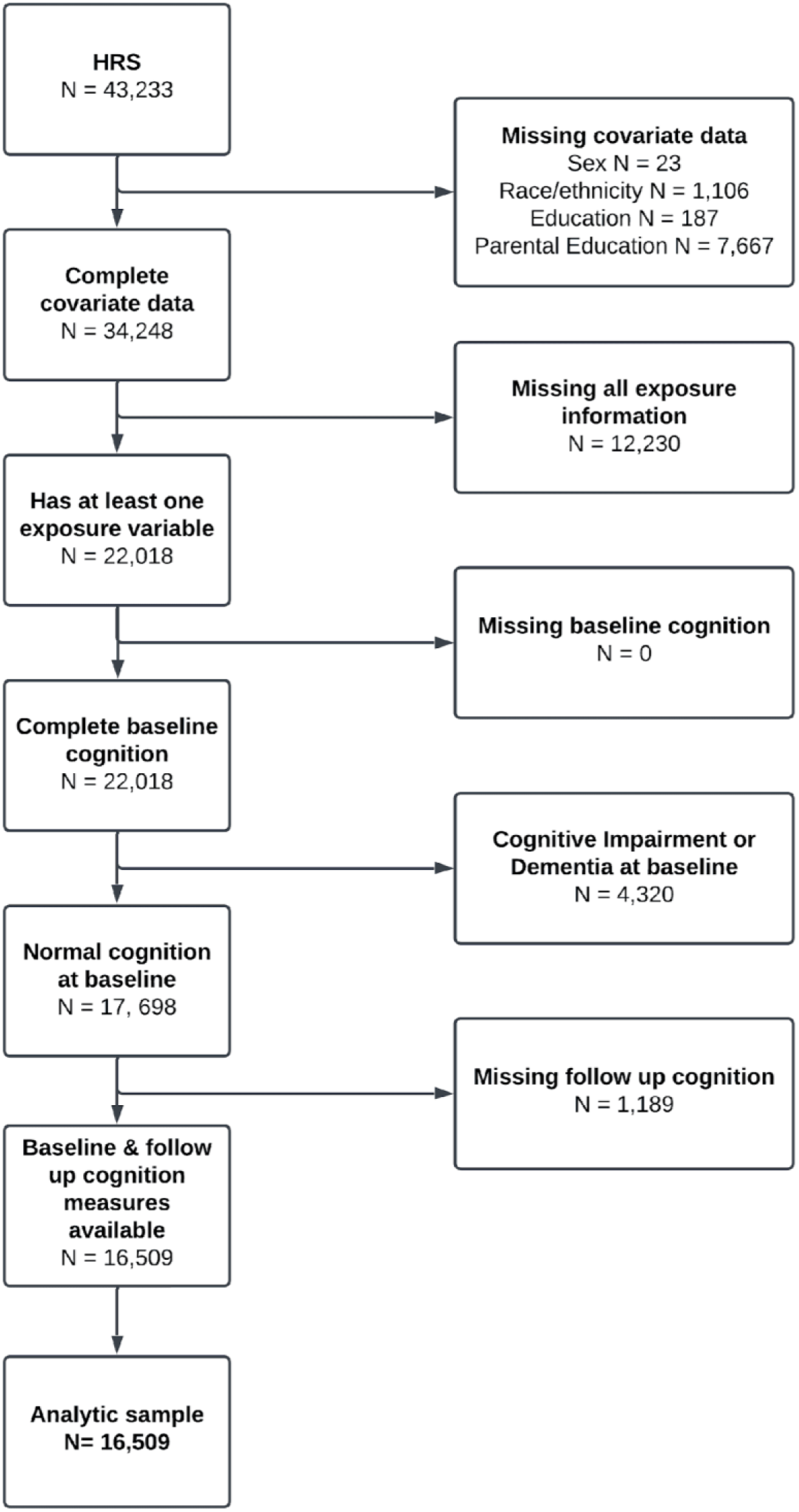
Flow chart illustrating study Individual selection from the Health and Retirement Study (HRS) waves 2008 to 2018. Exposure data (waves 2008-2014) and cognition data (waves 2008-2018) were obtained from the HRS for all Individual s. Those with incomplete data for sex, race/ethnicity, personal education, and parental education were excluded (n= 8,985). Individuals with data for at least one exposure variable and complete baseline cognition were eligible for the study. We further excluded individuals who presented with cognitively impaired non-dementia or dementia symptomatology at baseline (n= 4,320). Finally, we excluded individuals without at least one follow-up cognition visit (n=1,189).

**Figure 2.**
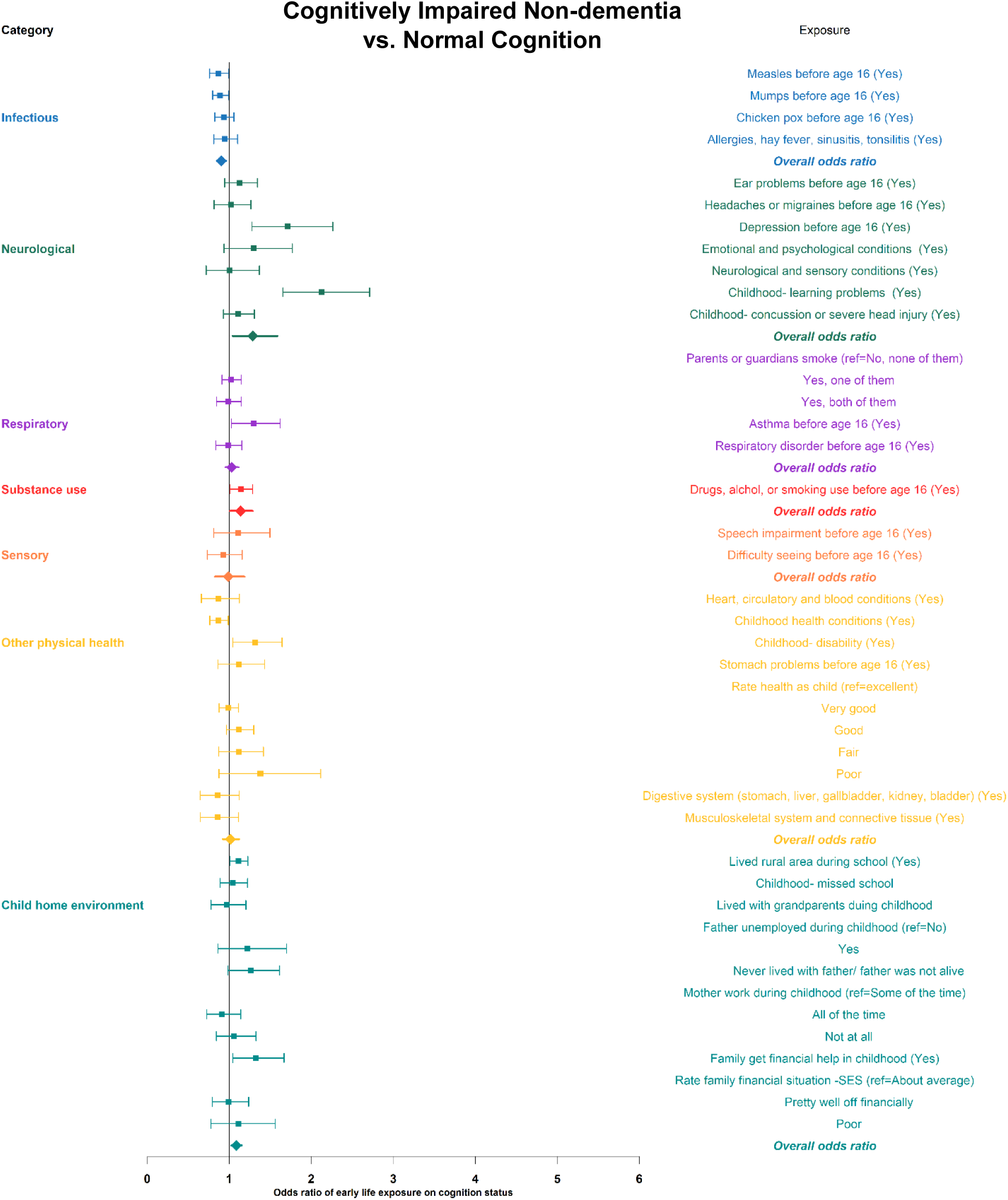
Forest plot displaying odds ratios for each early life exposure or condition (n=31) analyzed from HRS waves 2008-2014, comparing those with cognitive impairment non-dementia relative to normal cognition using cognition data from the 2008-2018 waves of the HRS on the same Individual s.

**Figure 3.**
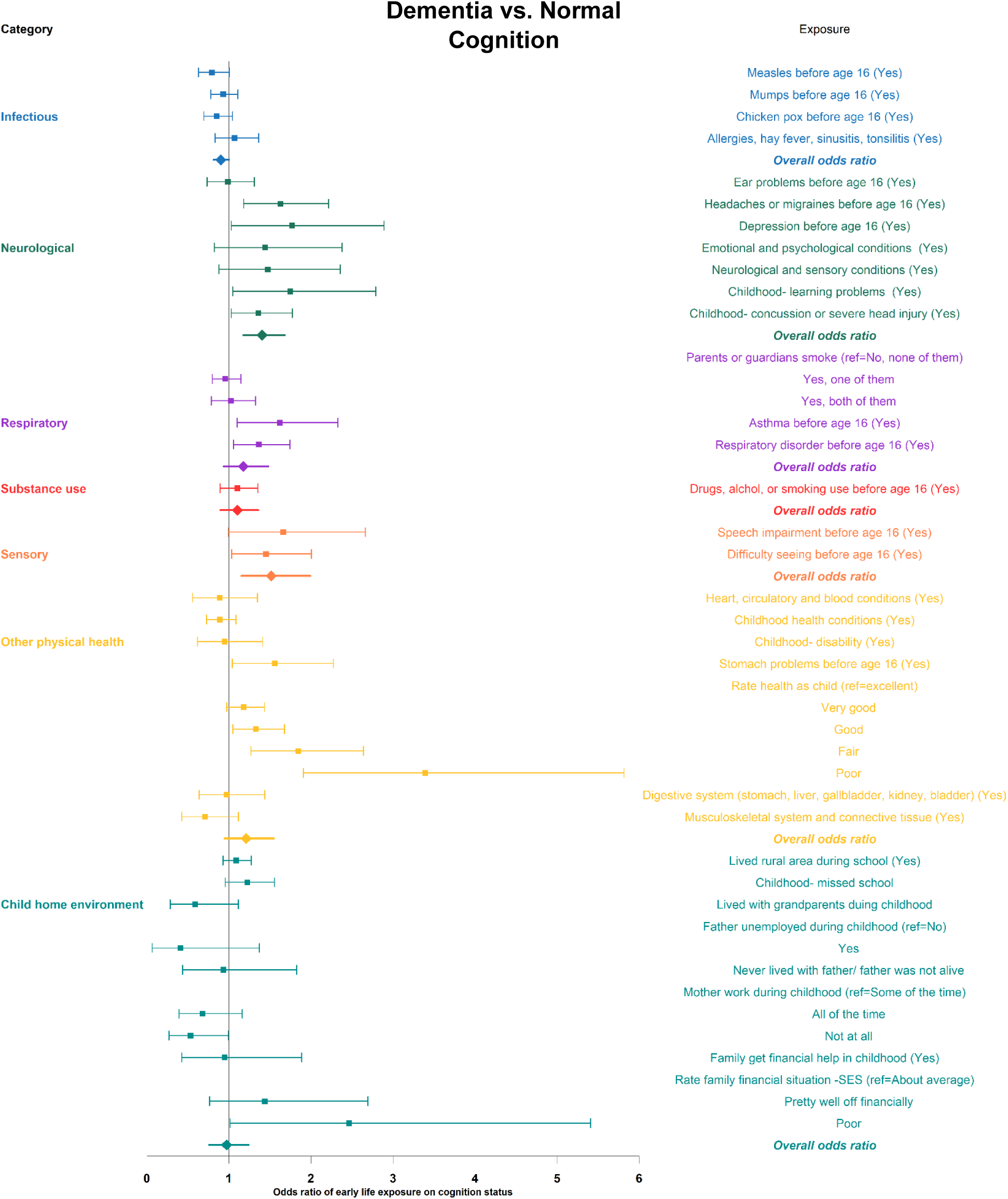
Forest plot displaying odds ratios for each early life exposure or condition (n=31) analyzed from HRS waves 2008-2014, comparing those with dementia relative to normal cognition using cognition data from the 2008-2018 waves of the HRS on the same Individual s. Exposure categories are

In the included sample at the end of follow-up, 13,236 (80%) Individuals presented with normal cognition, 2,395 (14.5%) had cognitive impairment non-dementia, and 878 (5.3%) had dementia (**Table 1**). Ten years of follow-up were completed for 42% of those with normal cognition, 38% with cognitive Impairment non-dementia, and 36% of those with dementia. We saw significant differences in the proportion of those exposed to allergies, measles, chicken pox, childhood learning problems, concussion or severe head injury, parent/guardian smoking, respiratory disorders, drugs, alcohol, or smoking use, childhood health conditions, self-rated health as a child, musculoskeletal and connective tissue issues, rural area of residence, self-rated family financial situation, paternal unemployment during childhood, maternal work during childhood, and family needed financial help in childhood (p-values<0.05) by cognitive status at follow-up. (**Table 1**).

**Table 1.** Bivariate Characteristics of Included Study Individual s in Health and Retirement Study (HRS) Sample 2008-2018.

|  |  |  |  |  |  |  |
| --- | --- | --- | --- | --- | --- | --- |
| Characteristic | N | Cognitive Status at Follow-up: |  |  |  | P-value<sup>2</sup> |
|  |  | Overall | Normal | Cognitively Impaired Non-Dementia | Dementia |  |
|  |  | N = 16,509 | N = 13,236 | N = 2,395 | N = 878 |  |
| **Time from baseline to end of follow up (years)** | 16,509 |  |  |  |  | <0.001 |
| 2 |  | 1,125 (6.8%) | 873 (6.6%) | 199 (8.3%) | 53 (6.0%) |  |
| 4 |  | 1,445 (8.8%) | 1,075 (8.1%) | 275 (11%) | 95 (11%) |  |
| 6 |  | 2,168 (13%) | 1,604 (12%) | 382 (16%) | 182 (21%) |  |
| 8 |  | 4,978 (30%) | 4,104 (31%) | 638 (27%) | 236 (27%) |  |
| 10 |  | 6,793 (41%) | 5,580 (42%) | 901 (38%) | 312 (36%) |  |
| **Age at baseline** | 16,509 | 63 (11) | 61 (10) | 68 (11) | 74 (10) | <0.001 |
| **Proxy status at baseline** | 16,509 |  |  |  |  | 0.4 |
| No proxy |  | 16,356 (99%) | 13,117 (99%) | 2,373 (99%) | 866 (99%) |  |
| Proxy |  | 153 (0.9%) | 119 (0.9%) | 22 (0.9%) | 12 (1.4%) |  |
| **Proxy status at follow-up** | 16,509 |  |  |  |  | <0.001 |
| No proxy |  | 15,697 (95%) | 13,006 (98%) | 2,282 (95%) | 409 (47%) |  |
| Proxy |  | 812 (4.9%) | 230 (1.7%) | 113 (4.7%) | 469 (53%) |  |
| **Personal educational attainment (years)** | 16,509 | 13.29 (2.75) | 13.55 (2.61) | 12.23 (2.99) | 12.29 (3.15) | <0.001 |
| **Sex** | 16,509 |  |  |  |  | 0.001 |
| Male |  | 6,737 (41%) | 5,372 (41%) | 1,042 (44%) | 323 (37%) |  |
| Female |  | 9,772 (59%) | 7,864 (59%) | 1,353 (56%) | 555 (63%) |  |
| **Race/ethnicity** | 16,509 |  |  |  |  | <0.001 |
| Non-Hispanic White |  | 11,617 (70%) | 9,413 (71%) | 1,546 (65%) | 658 (75%) |  |
| Non-Hispanic Black |  | 2,526 (15%) | 1,923 (15%) | 480 (20%) | 123 (14%) |  |
| Hispanic |  | 1,826 (11%) | 1,456 (11%) | 285 (12%) | 85 (9.7%) |  |
| Non-Hispanic Other |  | 540 (3.3%) | 444 (3.4%) | 84 (3.5%) | 12 (1.4%) |  |
| **Parental highest degree of education** | 16,509 |  |  |  |  | <0.001 |
| >= High School |  | 14,009 (85%) | 11,502 (87%) | 1,837 (77%) | 670 (76%) |  |
| < High School |  | 2,500 (15%) | 1,734 (13%) | 558 (23%) | 208 (24%) |  |
| **Exposures before age 16** |  |  |  |  |  |  |
| **Infectious** |  |  |  |  |  |  |
| **Allergies; hay fever; sinusitis; tonsillitis** | 16,328 |  |  |  |  | <0.001 |
| No |  | 14,241 (87%) | 11,334 (87%) | 2,136 (90%) | 771 (90%) |  |
| Yes |  | 2,087 (13%) | 1,763 (13%) | 234 (9.9%) | 90 (10%) |  |
| Missing |  | 181 | 139 | 25 | 17 |  |
| **Measles** | 12,530 |  |  |  |  | <0.001 |
| No |  | 2,808 (22%) | 2,284 (23%) | 402 (20%) | 122 (17%) |  |
| Yes |  | 9,722 (78%) | 7,532 (77%) | 1,590 (80%) | 600 (83%) |  |
| Missing |  | 3,979 | 3,420 | 403 | 156 |  |
| **Mumps** | 12,437 |  |  |  |  | 0.4 |
| No |  | 4,906 (39%) | 3,860 (40%) | 779 (40%) | 267 (37%) |  |
| Yes |  | 7,531 (61%) | 5,887 (60%) | 1,191 (60%) | 453 (63%) |  |
| Missing |  | 4,072 | 3,489 | 425 | 158 |  |
| **Chicken pox** | 12,494 |  |  |  |  | <0.001 |
| No |  | 2,735 (22%) | 2,052 (21%) | 504 (26%) | 179 (25%) |  |
| Yes |  | 9,759 (78%) | 7,761 (79%) | 1,464 (74%) | 534 (75%) |  |
| Missing |  | 4,015 | 3,423 | 427 | 165 |  |
| **Neurological** |  |  |  |  |  |  |
| **Ear problems** | 12,950 |  |  |  |  | >0.9 |
| No |  | 11,751 (91%) | 9,202 (91%) | 1,875 (91%) | 674 (91%) |  |
| Yes |  | 1,199 (9.3%) | 944 (9.3%) | 187 (9.1%) | 68 (9.2%) |  |
| Missing |  | 3,559 | 3,090 | 333 | 136 |  |
| **Headaches or migraines** | 12,956 |  |  |  |  | 0.055 |
| No |  | 12,132 (94%) | 9,496 (94%) | 1,951 (95%) | 685 (92%) |  |
| Yes |  | 824 (6.4%) | 654 (6.4%) | 112 (5.4%) | 58 (7.8%) |  |
| Missing |  | 3,553 | 3,086 | 332 | 135 |  |
| **Depression** | 12,951 |  |  |  |  | 0.7 |
| No |  | 12,540 (97%) | 9,822 (97%) | 1,996 (97%) | 722 (97%) |  |
| Yes |  | 411 (3.2%) | 323 (3.2%) | 68 (3.3%) | 20 (2.7%) |  |
| Missing |  | 3,558 | 3,091 | 331 | 136 |  |
| **Emotional and psychological conditions** | 16,322 |  |  |  |  | >0.9 |
| No |  | 15,950 (98%) | 12,790 (98%) | 2,318 (98%) | 842 (98%) |  |
| Yes |  | 372 (2.3%) | 301 (2.3%) | 52 (2.2%) | 19 (2.2%) |  |
| Missing |  | 187 | 145 | 25 | 17 |  |
| **Neurological and sensory conditions** | 13,025 |  |  |  |  | 0.2 |
| No |  | 12,635 (97%) | 9,893 (97%) | 2,018 (98%) | 724 (97%) |  |
| Yes |  | 390 (3.0%) | 320 (3.1%) | 49 (2.4%) | 21 (2.8%) |  |
| Missing |  | 3,484 | 3,023 | 328 | 133 |  |
| **Childhood learning problems** | 16,348 |  |  |  |  | <0.001 |
| No |  | 15,880 (97%) | 12,767 (97%) | 2,270 (96%) | 843 (97%) |  |
| Yes |  | 468 (2.9%) | 344 (2.6%) | 102 (4.3%) | 22 (2.5%) |  |
| Missing |  | 161 | 125 | 23 | 13 |  |
| **Concussion or severe head injury** | 16,325 |  |  |  |  | 0.017 |
| No |  | 14,778 (91%) | 11,807 (90%) | 2,180 (92%) | 791 (92%) |  |
| Yes |  | 1,547 (9.5%) | 1,284 (9.8%) | 190 (8.0%) | 73 (8.4%) |  |
| Missing |  | 184 | 145 | 25 | 14 |  |
| **Respiratory** |  |  |  |  |  |  |
| **Parents/guardians smoke** | 12,914 |  |  |  |  | <0.001 |
| No, none of them |  | 4,410 (34%) | 3,385 (33%) | 736 (36%) | 289 (39%) |  |
| Yes, one of them |  | 5,752 (45%) | 4,411 (44%) | 986 (48%) | 355 (48%) |  |
| Yes, both |  | 2,752 (21%) | 2,325 (23%) | 329 (16%) | 98 (13%) |  |
| Missing |  | 3,595 | 3,115 | 344 | 136 |  |
| **Asthma** | 12,954 |  |  |  |  | >0.9 |
| No |  | 12,277 (95%) | 9,618 (95%) | 1,955 (95%) | 704 (95%) |  |
| Yes |  | 677 (5.2%) | 531 (5.2%) | 107 (5.2%) | 39 (5.2%) |  |
| Missing |  | 3,555 | 3,087 | 333 | 135 |  |
| **Respiratory disorder** | 12,939 |  |  |  |  | <0.001 |
| No |  | 11,167 (86%) | 8,672 (86%) | 1,845 (89%) | 650 (87%) |  |
| Yes |  | 1,772 (14%) | 1,462 (14%) | 217 (11%) | 93 (13%) |  |
| Missing |  | 3,570 | 3,102 | 333 | 135 |  |
| **Substance use/Behavioral** |  |  |  |  |  |  |
| **Drugs, alcohol, or smoking use** | 13,621 |  |  |  |  | 0.036 |
| No |  | 10,485 (77%) | 8,215 (77%) | 1,647 (77%) | 623 (81%) |  |
| Yes |  | 3,136 (23%) | 2,486 (23%) | 501 (23%) | 149 (19%) |  |
| Missing |  | 2,888 | 2,535 | 247 | 106 |  |
| **Sensory** |  |  |  |  |  |  |
| **Difficulty seeing** | 12,958 |  |  |  |  | 0.12 |
| No |  | 12,230 (94%) | 9,576 (94%) | 1,961 (95%) | 693 (93%) |  |
| Yes |  | 728 (5.6%) | 576 (5.7%) | 101 (4.9%) | 51 (6.9%) |  |
| Missing |  | 3,551 | 3,084 | 333 | 134 |  |
| **Speech impairment** | 12,962 |  |  |  |  | 0.3 |
| No |  | 12,544 (97%) | 9,813 (97%) | 2,009 (97%) | 722 (97%) |  |
| Yes |  | 418 (3.2%) | 341 (3.4%) | 55 (2.7%) | 22 (3.0%) |  |
| Missing |  | 3,547 | 3,082 | 331 | 134 |  |
| **Other Physical Health** |  |  |  |  |  |  |
| **Heart, circulatory and blood conditions** | 13,009 |  |  |  |  | 0.11 |
| No |  | 12,462 (96%) | 9,754 (96%) | 1,993 (96%) | 715 (97%) |  |
| Yes |  | 547 (4.2%) | 449 (4.4%) | 73 (3.5%) | 25 (3.4%) |  |
| Missing |  | 3,500 | 3,033 | 329 | 138 |  |
| **Childhood health conditions** | 16,327 |  |  |  |  | 0.003 |
| No |  | 13,404 (82%) | 10,686 (82%) | 2,002 (85%) | 716 (83%) |  |
| Yes |  | 2,923 (18%) | 2,410 (18%) | 367 (15%) | 146 (17%) |  |
| Missing |  | 182 | 140 | 26 | 16 |  |
| **Childhood disability** | 16,344 |  |  |  |  | 0.072 |
| No |  | 15,728 (96%) | 12,632 (96%) | 2,263 (95%) | 833 (97%) |  |
| Yes |  | 616 (3.8%) | 477 (3.6%) | 109 (4.6%) | 30 (3.5%) |  |
| Missing |  | 165 | 127 | 23 | 15 |  |
| **Stomach problems** | 12,952 |  |  |  |  | 0.5 |
| No |  | 12,372 (96%) | 9,688 (95%) | 1,979 (96%) | 705 (95%) |  |
| Yes |  | 580 (4.5%) | 459 (4.5%) | 84 (4.1%) | 37 (5.0%) |  |
| Missing |  | 3,557 | 3,089 | 332 | 136 |  |
| **Rate health as child** | 12,908 |  |  |  |  | <0.001 |
| Excellent |  | 6,720 (52%) | 5,413 (54%) | 1,002 (49%) | 305 (41%) |  |
| Very Good |  | 3,602 (28%) | 2,785 (28%) | 584 (28%) | 233 (31%) |  |
| Good |  | 1,813 (14%) | 1,336 (13%) | 337 (16%) | 140 (19%) |  |
| Fair |  | 609 (4.7%) | 459 (4.5%) | 104 (5.1%) | 46 (6.2%) |  |
| Poor |  | 164 (1.3%) | 113 (1.1%) | 30 (1.5%) | 21 (2.8%) |  |
| Missing |  | 3,601 | 3,130 | 338 | 133 |  |
| **Digestive system (stomach, liver, gallbladder, kidney, bladder)** | 16,322 |  |  |  |  | 0.2 |
| No |  | 15,761 (97%) | 12,627 (96%) | 2,303 (97%) | 831 (96%) |  |
| Yes |  | 561 (3.4%) | 464 (3.5%) | 66 (2.8%) | 31 (3.6%) |  |
| Missing |  | 187 | 145 | 26 | 16 |  |
| **Musculoskeletal system and connective tissue** | 16,321 |  |  |  |  | 0.014 |
| No |  | 15,730 (96%) | 12,590 (96%) | 2,299 (97%) | 841 (98%) |  |
| Yes |  | 591 (3.6%) | 501 (3.8%) | 70 (3.0%) | 20 (2.3%) |  |
| Missing |  | 188 | 145 | 26 | 17 |  |
| **Child Home Environment** |  |  |  |  |  |  |
| **Lived in rural area as a child** | 16,132 |  |  |  |  | <0.001 |
| No |  | 9,322 (58%) | 7,641 (59%) | 1,225 (53%) | 456 (55%) |  |
| Yes |  | 6,810 (42%) | 5,357 (41%) | 1,084 (47%) | 369 (45%) |  |
| Missing |  | 377 | 238 | 86 | 53 |  |
| **Missed school as a child** | 12,930 |  |  |  |  | 0.15 |
| No |  | 11,473 (89%) | 9,005 (89%) | 1,825 (89%) | 643 (87%) |  |
| Yes |  | 1,457 (11%) | 1,122 (11%) | 235 (11%) | 100 (13%) |  |
| Missing |  | 3,579 | 3,109 | 335 | 135 |  |
| **Rate family financial situation** | 5,046 |  |  |  |  | <0.001 |
| About Avg |  | 3,111 (62%) | 2,822 (62%) | 266 (56%) | 23 (42%) |  |
| Poor |  | 1,410 (28%) | 1,220 (27%) | 166 (35%) | 24 (44%) |  |
| Pretty well off financially |  | 525 (10%) | 474 (10%) | 43 (9.1%) | 8 (15%) |  |
| Missing |  | 11,463 | 8,720 | 1,920 | 823 |  |
| **Live with grandparents during childhood** | 5,083 |  |  |  |  | 0.3 |
| No |  | 3,721 (73%) | 3,336 (73%) | 340 (71%) | 45 (80%) |  |
| Yes |  | 1,362 (27%) | 1,212 (27%) | 139 (29%) | 11 (20%) |  |
| Missing |  | 11,426 | 8,688 | 1,916 | 822 |  |
| **Father unemployed during childhood** | 5,024 |  |  |  |  | 0.002 |
| No |  | 3,808 (76%) | 3,438 (77%) | 327 (69%) | 43 (78%) |  |
| Never lived with father/ father was not alive |  |  | 291 (6.5%) | 50 (11%) | 2 (3.6%) |  |
| Yes |  | 873 (17%) | 764 (17%) | 99 (21%) | 10 (18%) |  |
| Missing |  | 11,485 | 8,743 | 1,919 | 823 |  |
| **Mother work during childhood** | 5,467 |  |  |  |  | 0.044 |
| Some of the time |  | 2,151 (39%) | 1,917 (40%) | 201 (36%) | 33 (40%) |  |
| Not at all |  | 1,842 (34%) | 1,618 (34%) | 189 (34%) | 35 (43%) |  |
| All of the time |  | 1,474 (27%) | 1,290 (27%) | 170 (30%) | 14 (17%) |  |
| Missing |  | 11,042 | 8,411 | 1,835 | 796 |  |
| **Family get financial help in childhood** | 4,986 |  |  |  |  | 0.022 |
| No |  | 4,008 (80%) | 3,607 (81%) | 355 (76%) | 46 (84%) |  |
| Yes |  | 978 (20%) | 855 (19%) | 114 (24%) | 9 (16%) |  |
| Missing |  | 11,523 | 8,774 | 1,926 | 823 |  |

### Exposure associations with incident cognitive impairment non-dementia

Crude model results are available (**Supplemental Table 3**). After covariate adjustment, two exposures were associated with cognitive impairment non-dementia status relative to normal cognition using our stringent criteria (FDR<0.05) (**Table 2**). Having depression before the age of 16 years old was associated with 1.71 (95% CI: 1.28, 2.26) times increased odds of incident cognitive impairment non-dementia at follow-up. The presence of childhood learning problems before the age of 16 years old was associated with 2.12 (95% CI: 1.65, 2.71) times increased odds of incident cognitive impairment non-dementia during follow-up.

**Table 2.** Adjusted Logistic Regression of Incident Cognition and Exposures or Conditions before age 16 in the Health and Retirement Study (HRS) 2008-2018.

| Dementia vs Cognitively normal |  |  |  |  |  |  | Cognitively impaired non-dementia<br>vs Cognitively normal |  |  |  |
| --- | --- | --- | --- | --- | --- | --- | --- | --- | --- | --- |
| Exposures before age 16 | Exposed | Responded | OR | (95% CI) | P-value | FDR | OR | (95% CI) | P-value | FDR |
| Infectious |  |  |  |  |  |  |  |  |  |  |
| Measles | 9,722 | 12,530 | 0.793 | (0.629, 1.004) | 0.051 | 0.138 | 0.867 | (0.759, 0.992) | 0.037 | 0.119 |
| Mumps | 7,531 | 12,437 | 0.928 | (0.778, 1.108) | 0.407 | 0.523 | 0.888 | (0.797, 0.99) | 0.031 | 0.119 |
| Chicken pox | 9,759 | 12,494 | 0.849 | (0.693, 1.044) | 0.117 | 0.243 | 0.931 | (0.822, 1.056) | 0.262 | 0.510 |
| Allergies, hay fever, sinusitis, tonsillitis | 2,087 | 16,328 | 1.07 | (0.832, 1.361) | 0.591 | 0.719 | 0.947 | (0.811, 1.101) | 0.483 | 0.714 |
| Overall odds ratio |  |  | 0.901 | (0.811, 1.001) | 0.052 | 0.138 | 0.904 | (0.848, 0.963) | 0.002 | 0.030 |
| Neurological |  |  |  |  |  |  |  |  |  |  |
| Headaches or migraines | 824 | 12,956 | 1.629 | (1.18, 2.215) | 0.002 | 0.022 | 1.018 | (0.814, 1.263) | 0.875 | 0.929 |
| Ear problems | 1,199 | 12,950 | 0.987 | (0.734, 1.309) | 0.930 | 0.930 | 1.127 | (0.942, 1.343) | 0.186 | 0.441 |
| Depression | 411 | 12,951 | 1.769 | (1.03, 2.89) | 0.030 | 0.104 | 1.709 | (1.276, 2.261) | 0.000 | 0.000 |
| Emotional and psychological conditions | 372 | 16,322 | 1.439 | (0.823, 2.38) | 0.177 | 0.284 | 1.299 | (0.936, 1.769) | 0.107 | 0.301 |
| Neurological and sensory conditions | 390 | 13,025 | 1.473 | (0.878, 2.355) | 0.122 | 0.243 | 1 | (0.717, 1.366) | 0.998 | 0.998 |
| Childhood learning problems | 468 | 16,348 | 1.748 | (1.046, 2.788) | 0.025 | 0.101 | 2.123 | (1.652, 2.71) | 0.000 | 0.000 |
| Childhood concussion/severe head injury | 1,547 | 16,325 | 1.359 | (1.028, 1.773) | 0.027 | 0.101 | 1.104 | (0.929, 1.306) | 0.255 | 0.510 |
| Overall odds ratio |  |  | 1.404 | (1.174, 1.681) | 0.000 | 0.000 | 1.285 | (1.041, 1.587) | 0.020 | 0.119 |
| Respiratory |  |  |  |  |  |  |  |  |  |  |
| Parents or guardians smoke<sup>1</sup> |  | 12,914 |  |  |  |  |  |  |  |  |
| Yes, one of them | 5,752 |  | 0.956 | (0.797, 1.148) | 0.630 | 0.746 | 1.022 | (0.911, 1.145) | 0.715 | 0.894 |
| Yes, both | 2,752 |  | 1.024 | (0.786, 1.325) | 0.861 | 0.916 | 0.983 | (0.843, 1.144) | 0.821 | 0.919 |
| Asthma | 677 | 12,954 | 1.62 | (1.1, 2.328) | 0.012 | 0.090 | 1.294 | (1.025, 1.62) | 0.027 | 0.119 |
| Respiratory disorder | 1,772 | 12,939 | 1.363 | (1.057, 1.743) | 0.015 | 0.096 | 0.983 | (0.835, 1.152) | 0.833 | 0.919 |
| Overall odds ratio |  |  | 1.177 | (0.936, 1.481) | 0.164 | 0.273 | 1.029 | (0.955, 1.11) | 0.449 | 0.697 |
| Behavioral/substance use |  |  |  |  |  |  |  |  |  |  |
| Drugs, alcohol, or smoking use | 3,136 | 13,621 | 1.102 | (0.893, 1.353) | 0.361 | 0.478 | 1.138 | (1.008, 1.284) | 0.036 | 0.119 |
| Sensory |  |  |  |  |  |  |  |  |  |  |
| Speech impairment | 418 | 12,962 | 1.661 | (0.991, 2.661) | 0.043 | 0.129 | 1.109 | (0.809, 1.494) | 0.508 | 0.714 |
| Difficulty seeing | 728 | 12,958 | 1.452 | (1.033, 2.005) | 0.027 | 0.101 | 0.924 | (0.73, 1.158) | 0.499 | 0.714 |
| Overall odds ratio |  |  | 1.514 | (1.151, 1.993) | 0.003 | 0.027 | 0.987 | (0.821, 1.186) | 0.888 | 0.929 |
| Other Physical Health |  |  |  |  |  |  |  |  |  |  |
| Heart, circulatory and blood conditions | 547 | 13,009 | 0.886 | (0.559, 1.347) | 0.590 | 0.719 | 0.866 | (0.659, 1.122) | 0.288 | 0.518 |
| Childhood health conditions | 2,923 | 16,327 | 0.89 | (0.725, 1.085) | 0.255 | 0.373 | 0.868 | (0.763, 0.986) | 0.030 | 0.119 |
| Childhood disability | 616 | 16,344 | 0.949 | (0.617, 1.411) | 0.805 | 0.916 | 1.314 | (1.043, 1.643) | 0.018 | 0.119 |
| Stomach problems | 580 | 12,952 | 1.557 | (1.04, 2.271) | 0.026 | 0.101 | 1.115 | (0.86, 1.431) | 0.400 | 0.653 |
| Rate health as child <sup>2</sup> |  | 12,908 |  |  |  |  |  |  |  |  |
| Very good | 3,602 |  | 1.181 | (0.972, 1.434) | 0.093 | 0.220 | 0.988 | (0.876, 1.113) | 0.837 | 0.919 |
| Good | 1,813 |  | 1.329 | (1.048, 1.678) | 0.018 | 0.101 | 1.119 | (0.964, 1.298) | 0.137 | 0.363 |
| Fair | 609 |  | 1.845 | (1.267, 2.64) | 0.001 | 0.015 | 1.115 | (0.872, 1.415) | 0.377 | 0.653 |
| Poor | 164 |  | 3.392 | (1.906, 5.815) | 0.000 | 0.000 | 1.379 | (0.874, 2.114) | 0.153 | 0.382 |
| Digestive system (stomach, liver, gallbladder, kidney, bladder) | 561 | 16,322 | 0.973 | (0.637, 1.438) | 0.896 | 0.916 | 0.857 | (0.645, 1.121) | 0.272 | 0.510 |
| Musculoskeletal system and connective tissue | 591 | 16,321 | 0.706 | (0.424, 1.115) | 0.157 | 0.273 | 0.854 | (0.647, 1.112) | 0.254 | 0.510 |
| **Overall odds ratio** |  |  | 1.21 | (0.945, 1.55) | 0.131 | 0.246 | 1.015 | (0.92, 1.119) | 0.771 | 0.919 |
| **Child Home Environment** |  |  |  |  |  |  |  |  |  |  |
| Lived rural area during school | 6,810 | 16,132 | 1.088 | (0.929, 1.273) | 0.297 | 0.418 | 1.112 | (1.009, 1.225) | 0.032 | 0.119 |
| Childhood Missed school | 1,457 | 12,930 | 1.224 | (0.956, 1.555) | 0.102 | 0.229 | 1.043 | (0.888, 1.222) | 0.604 | 0.799 |
| Live with grandparents during childhood | 1,362 | 5,083 | 0.588 | (0.285, 1.116) | 0.124 | 0.243 | 0.969 | (0.778, 1.202) | 0.777 | 0.919 |
| Father unemployed during childhood <sup>3</sup> |  | 5,024 |  |  |  |  |  |  |  |  |
| Never lived with father/ father not alive | 343 |  | 0.409 | (0.066, 1.371) | 0.224 | 0.348 | 1.219 | (0.86, 1.698) | 0.253 | 0.510 |
| Very good | 873 |  | 0.934 | (0.434, 1.825) | 0.851 | 0.916 | 1.264 | (0.982, 1.614) | 0.064 | 0.192 |
| Mother work during childhood <sup>4</sup> | 3,316 | 5,467 |  |  |  |  |  |  |  |  |
| Not at all | 1,842 |  | 0.677 | (0.392, 1.162) | 0.158 | 0.273 | 0.908 | (0.722, 1.14) | 0.406 | 0.653 |
| All of the time | 1,474 |  | 0.531 | (0.27, 0.992) | 0.055 | 0.138 | 1.057 | (0.842, 1.324) | 0.632 | 0.813 |
| Family get financial help in childhood | 978 | 4,986 | 0.947 | (0.426, 1.886) | 0.884 | 0.916 | 1.323 | (1.044, 1.667) | 0.019 | 0.119 |
| Rate family financial situation<sup>5</sup> | 1,935 | 5,046 |  |  |  |  |  |  |  |  |
| Poor | 1,410 |  | 1.437 | (0.764, 2.693) | 0.257 | 0.373 | 0.991 | (0.791, 1.237) | 0.933 | 0.954 |
| Pretty well off financially | 525 |  | 2.462 | (1.013, 5.408) | 0.033 | 0.106 | 1.112 | (0.776, 1.56) | 0.550 | 0.750 |
| **Overall odds ratio** |  |  | 0.971 | (0.76, 1.241) | 0.816 | 0.916 | 1.086 | (1.023, 1.153) | 0.007 | 0.079 |

HRS: Health and Retirement Study; CIND: Cognitive impairment Non-Dementia; OR: Odds Ratio; BP: Blood Pressure; SES: Socioeconomic status;
<sup>1</sup>Reference group in parents or guardians smoke is "No, None of them"; <sup>2</sup>Reference group in Rate health as child is "Excellent"; <sup>3</sup>Reference group in Father unemployed during childhood is "No"; <sup>4</sup>Reference group in Mother work during childhood is "Some of the time"; <sup>5</sup>Reference group in Rate family financial situation is "About average". Unless otherwise stated reference group used for binary (yes/no) variables is "no".
[1] Cognitive status was defined as the cognition status of the Individual at their most recent follow-up cognitive assessment visit (CIND, Dementia, or normal) [2] Exposures were retrospectively analyzed from a self-reported questionnaire completed once in the 2008-2014 waves of the HRS. [3] Two additional years of HRS cognitive data on the same Individual s were included (2016 & 2018 HRS waves cognitive measures only). With the addition of the 2016 and 2018 cognition measures, our cognitive data spans from 2008 to 2018. [4] "Responded" is the total number of people with a valid measurement record for that exposure. "Exposed" is the count of people who responded yes to that exposure. [5] Sex, Race Ethnicity, Age at baseline, Highest parental education, Highest personal education, and time from exposure to cognition measure, were adjusted for in each model [6] The P-value was calculated using logistic regression and interpreted as differences between groups [7] Random-effect meta-analyses was used to estimate the pooled odds ratios from all exposures under the same category.

Additionally, with our permissive criteria (p-value<0.05), five exposures were associated with cognitive impairment non-dementia status. Having measles was associated with 0.87 (95% CI: 0.76, 0.99) times lower odds of incident cognitive impairment non-dementia at follow-up. Similarly, having mumps was associated with 0.89 (95% CI: 0.80, 0.99) times lower odds of cognitive impairment non-dementia at follow-up. Conversely, asthma before the age of 16 years old was associated with 1.14 (95% CI: 1.03, 1.62) times higher odds of cognitive impairment non-dementia. The use of drugs, alcohol, or smoking before the age of 16 was associated with 1.14 (95% CI: 1.01, 1.28) times higher odds of cognitive impairment non-dementia in later life. Finally, living in a rural area before the age of 16 was associated with 1.11 (95% CI: 1.01, 1.23) times higher odds of incident cognitive impairment non-dementia at follow-up.

When summarizing associations within an exposure category, one category met our stringent criteria. Having exposure to any of the infectious agents was associated with 0.90 (95% CI: 0.85, 0.96) times lower odds of incident cognitive impairment non-dementia at follow-up.

### Exposure associations with incident dementia

Results for the crude model can be found in **Supplemental Table 3.** After adjustment for covariates, two exposure variables were associated with dementia status relative to normal cognition following our stringent criteria (FDR<0.05, **Table 2**). Having headaches or migraines before the age of 16 years old was associated with 1.63 (95% CI:1.18, 2.12) times increased odds of incident dementia over follow-up. Self-reported fair overall health before age 16 was associated with a 1.85 (95% CI:1.27, 2.64) times increased odds of incident dementia, compared to those with self-rated excellent health at follow-up. Likewise, self-reported poor overall health before age 16 was observed to have a 3.39 (95% CI:1.91, 5.81) times increased odds of incident dementia, compared to those with self-rated excellent health at follow-up.

Subsequently, using our permissive criteria (p-value<0.05) six exposures were associated with dementia. For exposures before age 16, living with depression 1.77 (95% CI:1.03, 2.89), facing learning problems 1.75 (95% CI:1.05, 2.79), undergoing a concussion or severe head injury 1.36 (95% CI:1.03,1.77), having a respiratory condition of asthma 1.62 (95% CI:1.1,2.33), experiencing any respiratory disorder 1.36 (95% CI:1.06, 1.74), or experiencing stomach problems 1.56 (95% CI:1.04, 2.7) were all associated with increased odds of incident dementia over follow-up.

When looking at overall associations within exposure categories, two categories met stringent criteria. Neurological conditions were associated with 1.4 (95% CI:1.17, 1.68) times increased odds of incident dementia relative to normal cognition over follow-up. Also, sensory conditions were associated with 1.51 (95% CI:1.15, 1.99) times increased odds of incident dementia relative to normal cognition at follow-up.

### Sensitivity analyses: Associations with any cognitive impairment

To increase statistical power, we combined outcomes of dementia with cognitive impairment non-dementia and tested associations between exposures and the new any cognitive impairment variable. Our findings for individual exposures were generally consistent with the primary analyses (**Supplemental Table 3).** Following our stringent criteria (FDR<0.05), adjusting for key covariates, four exposures (depression, learning problems, asthma, and poor self-reported health) were associated with any cognitive impairment relative to normal cognition.

Furthermore, two exposure categories (FDR<0.05) were associated with incident any cognitive impairment. Exposure to any neurologic conditions before age 16 was associated with 1.33 (95% CI: 1.10, 1.62) times increased odds of incident any cognitive impairment over follow-up. Conversely, having exposure to any infectious condition before age 16 was associated with 0.90 (95% CI: 0.85, 0.96) times lower odds of incident any cognitive impairment over follow-up.

## Discussion

To broadly screen the relationships between exposures and conditions in the early life period and incident dementia or cognitive impairment non-dementia status in later life, we conducted an exposure-wide association study in a large and diverse sample of older adults. In the 2008-2018 waves of the HRS we analyzed the associations between retrospective assessments of exposures before 16 with incident cognitive status over 10 years of follow-up. We found depression and childhood learning problems had the strongest increased odds for incident cognitive impairment non-dementia. Headaches or migraines, neurologic disorders, self-rated health of fair or poor, and sensory conditions had the strongest associations with incident dementia. Our hypothesis-generating findings illustrate that exposures and environments in early life may play an important role in determining cognition status later and prioritizing conditions for examination in future studies.

### Neurologic conditions

In adulthood, neurological conditions such as learning disorders, migraines, and depression are risk factors for Alzheimer’s and other dementias.^22–29^ A critical review of epidemiology studies on a subset of early life factors (genotype, learning disability, education, socioeconomic status, and body size) with Alzheimer’s disease found that learning disorders in early life are associated with later risk of Alzheimer’s and dementia.^23^ Similarly, our study suggests that learning problems in early life impact the incidence of both dementia and cognitive impairment non-dementia. Specific factors such as idea density, self-rated below-average performance in school, dyslexia, speech difficulties, and trouble reading have been associated with Alzheimer’s or dementia later in life.^22^ Atypical development or alterations in the language centers of the brain may lead to selective vulnerability in the language cortex, resulting in neurodegeneration in this same region later in life.^22,23^ Two cohort studies of 198 and 8,523 individuals further posit the anatomical location of neurological changes in childhood influences the susceptibility for cognitive impairment in the same location later in life, rather than producing an effect in another area of the brain.^22,25^ Therefore, disparities in access to quality education and educational services may also contribute to disparities in dementia incidence.^1,2^ Future research can build on these findings to determine how genetic and environmental factors interact to influence development, and how intermediate stages of cognitive decline such as mild cognitive impairment fit in the causal pathway.^23^ When taken together, educational difficulties in childhood may be antecedent risk factors for dementia or Alzheimer’s in late life.

Concerning migraines, a retrospective cohort study of 33,468 adults observed individuals with migraines had a 1.33-fold increased risk of Alzheimer’s and dementia, compared to those without migraines.^28^ In addition, a meta-analysis of six studies encompassing 291,549 adult individuals reports a pooled risk ratio of 1.28 (95% CI: 0.64,2.54) increase in risk associated with a history of migraine and having dementia.^29^ While the biological mechanism linking migraines or chronic headaches to dementia development is unknown, the overlap between pain and memory centers in the brain may provide a potential explanation.^29^ Pain centers in the brain serve an important role in memory, and those with chronic headaches or migraines have been observed to have reduced gray-matter volume in the structures involved in memory.^29^ Similarly, our study found that the presence of migraines was associated with increased odds of having dementia. However, our study is the first to investigate how migraines and headaches in early life may impact cognitive status in adulthood using ten years of later-life cognitive follow-up data.

Depression is strongly associated with the development of dementia in adults.^26^ Our results show that depression before age 16 may also play a role in cognitive status. Specifically, depression was associated with increased odds of having cognitive impairment non-dementia, and any cognitive impairment via sensitivity analysis. Biological mechanisms such as reduced cerebral blood flow, chronic inflammation, and reduced serotonin availability may contribute to this association.^26,27^ A case-control study from 2020 examined 62,317 adult individuals from the German Disease Analyzer database and observed that anti-depressant treatment reduced the risk of incident dementia.^26^ Increasing the availability of serotonin in the brain through anti-depressant treatment may reduce the burden of amyloid plaques within the brain and allow for improved cognition.^26^ More studies investigating psychological factors in adolescence may contribute to identifying dementia at-risk humans as early as possible.

### Respiratory conditions

A meta-analysis of 15 studies with adult and child Individuals found cognitive impairments were associated with asthma.^30^ In particular, brain regions responsible for functions like executive functioning, language, learning, and memory had the strongest associations.^12,30^ During childhood, the brain is especially vulnerable to changes outside of homeostasis; In cases of asthma attacks, oxygen availability can fall below normal levels, which may lead to developmental delays, neurotransmitter imbalance, and neuronal cell death.^30–33^ These multifunctional brain regions have been implicated in the present study and current literature to have associations with previously identified risk factors such as learning disorders and migraines.^22,30^ However, researchers could not determine if asthma impacts the trajectory of normal cognitive aging outside of what is expected.^30^ Highlighting the need for studies investigating the impact of asthma on cognitive impairment across the life course specifically, in regions of the brain controlling executive function, academic achievement, learning, and memory which have been linked to dementia risk.^1–8,8,9,11–13,16,19,23–29,34,35^

### Infectious agents

We observed infectious diseases such as measles, and mumps were associated with reduced odds of cognitive impairment after follow-up. Similar to our findings, a cohort study of 2,994 individuals 65 and older from the Irish Longitudinal Study on Ageing found childhood infection with measles, mumps, or chickenpox, was associated with improved cognitive functioning later in life, and infection with all three was associated with increased cognitive functioning (β=0.18, 95% CI: 0.11, 0.26).^35^ The observed results in both of these studies are likely impacted by survival bias or unaccounted-for vaccination history, which may influence results. Potentially, residual antibodies produced during childhood infection may offer cognitive protection later in life.^35–37^ Antibodies produced after vaccination and mild stimulation of the immune system are hypothesized to have a protective effect on cognition in later life.^35,36,38^ Other studies have observed certain vaccinations were associated with reduced dementia risk and better cognitive function later in life.^37,38^ Overall, leveraging current vaccination systems may help curb the burden of dementia.

### Other childhood health conditions

This study and previous research in children and adults demonstrated a relationship between stomach health and dementia risk.^38–41^ A study of 200 Israeli Arab children, observed an association between *Helicobacter pylori* (*H. Pylori*) infection and lower overall cognition ability score.^40^ Intestinal infections like *H. Pylori* can result in intestinal malabsorption contributing to the development of iron deficiency anemia.^32,40^ Symptoms of iron deficiency anemia in children include decreased availability of oxygen within the body, a decline in school performance, and reduced learning capacity.^32,40^ These symptoms mirror other risk factors identified for dementia risk and future studies examining the role of stomach health as a moderating factor for dementia risk are needed.^27,30,31,33,37–40^ Furthermore, in adults, simultaneous infection of *H. pylori* and gum disease pathogens were observed to act synergistically to increase the risk of incident all-cause dementia in a study of adults 65 and older from the National Health and Nutrition Examination Survey (NHANES).^39^ Two mechanisms may explain this association. First, dysregulation of gut microbiota increases cytokine production and increases inflammation, which may activate microglia and deposit beta-amyloid within the brain.^38^ Second, the vagus nerve, a communication bridge between the brain, heart, and digestive system, is sensitive to changes in gut homeostasis.^38^ In animal studies, disruption of the vagus nerve was observed to elicit neurotransmitter imbalances in the brain similar to dementia (GABA, DOPA, glutamine).^33,38^ The relationship between dementia and the digestive system is complex, and future research on the role of stomach health on the causal pathway is needed.

### Limitations and future directions

Epidemiologic dementia research faces many methodological challenges.^42^ Because our exposures occurred before the study baseline and the recruitment age for the HRS begins at age 50, there is possible survival bias and selection bias that was not explicitly addressed in this study. Specifically, infectious exposures are subject to these biases as they are associated with mortality. Reliability of exposure assessment is subject to recall bias and is of particular concern when assessing those experiencing cognitive impairment or dementia. To minimize recall bias, we excluded individuals with cognitive impairment non-dementia or dementia at baseline and those missing all follow-up cognition information. In addition, because minoritized groups experience dementia at younger ages, excluding Individuals with cognitive impairment at baseline reduced the diversity of the included study sample. Dementia incidence is projected to disproportionally impact Black and Hispanic populations and studies comprehensively assessing exposures in these groups are of high public health importance.^1,2^

Our primary goal was to screen a wide variety of childhood exposures therefore we used an ExWAS data analysis framework, which models exposures independently and allows us to maximize the sample size for each exposure. Future studies that extend these discovery analyses by incorporating exposure mixtures or clustering analysis would be beneficial.^43^ It is important to note that many exposure mixtures analyses require complete (non-missing) observations. In our study, only a small subset (n=4,205) of our total (n=16,509) participants had responses to all 31 exposure variables, so these approaches may be challenging. Additionally, the HRS grouped these exposures into exposure summary categories based on the conceptual understanding of the study team. Future studies using data learning methods may give insight into exposure covariance structure, variance, and ensure the creation of the summary exposure categories are data driven.^44^

Consistent with the ExWAS framework we used a common model across exposures with standard covariates. Using these findings, analyses can follow-up with more extensive and customized analyses for individual exposures. For example, with some exposures, including health behavior variables such as BMI and smoking, or prevalent comorbidities such as adult-onset diabetes as confounder variables may be advantageous. For other exposures, these acquired behaviors or conditions in adulthood may either interact with exposures in early life, or be on the causal pathway to incident dementia, and may need to be modeled as effect modifiers or mediators. In addition, follow-up studies could include adjustment for polygenic scores for the specific exposure of interest, such as educational attainment or asthma, which may help control for the heritable component of the factor. Finally, replication testing is paramount to determine how childhood exposures fit in the causal pathway for disease development.

### Summary and implications

In the large and diverse HRS, this study screened 31 exposures before age 16 for association with incident dementia or cognitive impairment. Adjusting for age, sex, race, follow-up time, and education, 14 of the early life factors were associated with incident cognitive status. These findings broadly suggest that supporting growth and development in early life is critical for health throughout the life course. Alzheimer’s disease and other dementias pose a significant and growing public health burden globally. These findings suggest the early life period may be a relevant window of susceptibility and warrant more research. With further investigation prevention methods may be identified and implemented in future generations.

## Data Availability

The data presented in this study were accessed from the http://hrs.isr.umich.edu/data-products data repository: Public survey data: Cross-Wave Imputation of Cognitive Functioning Measures 1992-2020, release date May 2023 (Final V3.0); 2008 HRS Core release date December 2014 (Final V3.0); 2010 HRS Core release date August 2021 (Final V6.0); 2012 HRS Core release date March 2020 (Final V3.0); 2014 HRS Core release date December 2017 (Final V2.0); 2016 HRS Core release date December 2019 (Final V2.0).

https://hrsdata.isr.umich.edu/

http://hrs.isr.umich.edu/data-products

## Acknowledgments/conflicts/funding sources

We appreciate the Individuals and staff of the Health and Retirement Study. The Health and Retirement Study was supported by the National Institute on Aging (NIA U01AG009740). This analysis was supported by the National Institute on Aging (R01 AG067592, R01 AG070897, R01 AG055406, P30 AG072931). The authors gratefully acknowledge the use of the services and facilities of the Population Studies Center at the University of Michigan, funded by NICHD Center Grant P2CHD041028.

## Acronyms

CI: Confidence interval
ExWAS: Exposure-wide association framework
FDR: False discovery rate
HRS: Health and Retirement Study
NHANES: National Health and Nutrition Examination Survey
OR: Odds ratio
SD: Standard deviation

---

**Supplementary Figure 1.**
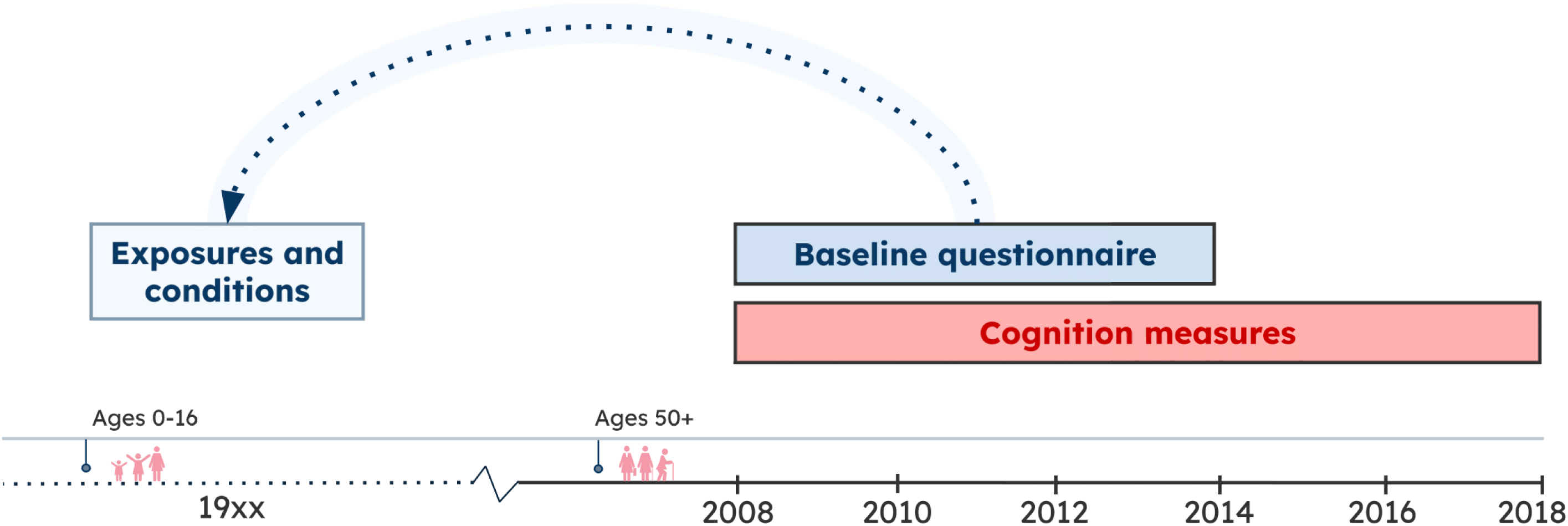
Timeline representation of the Health and Retirement Study (HRS) waves (2008-2018) used for this analysis. The HRS conducts biennial interviews with US adults 50 and older. Starting in 2008, 31 questions were incorporated into the core HRS questionnaire to retrospectively collect data on exposures and conditions before age 16. This one-time retrospective assessment includes eight exposure categories of infectious, neurologic, emotional and psychological, respiratory, sensory, substance use, physical health, and child home environment factors. Participants who did not respond to the 2008 early life exposure questionnaire were invited to respond at each subsequent visit. Exposure assessments completed between 2008 and 2014 were used as our baseline exposure assessment. Cognition tests are conducted at each follow-up wave, and this analysis used cognition measures from the 2008-2018 study waves on the same participants.

**Supplemental Table 1.**
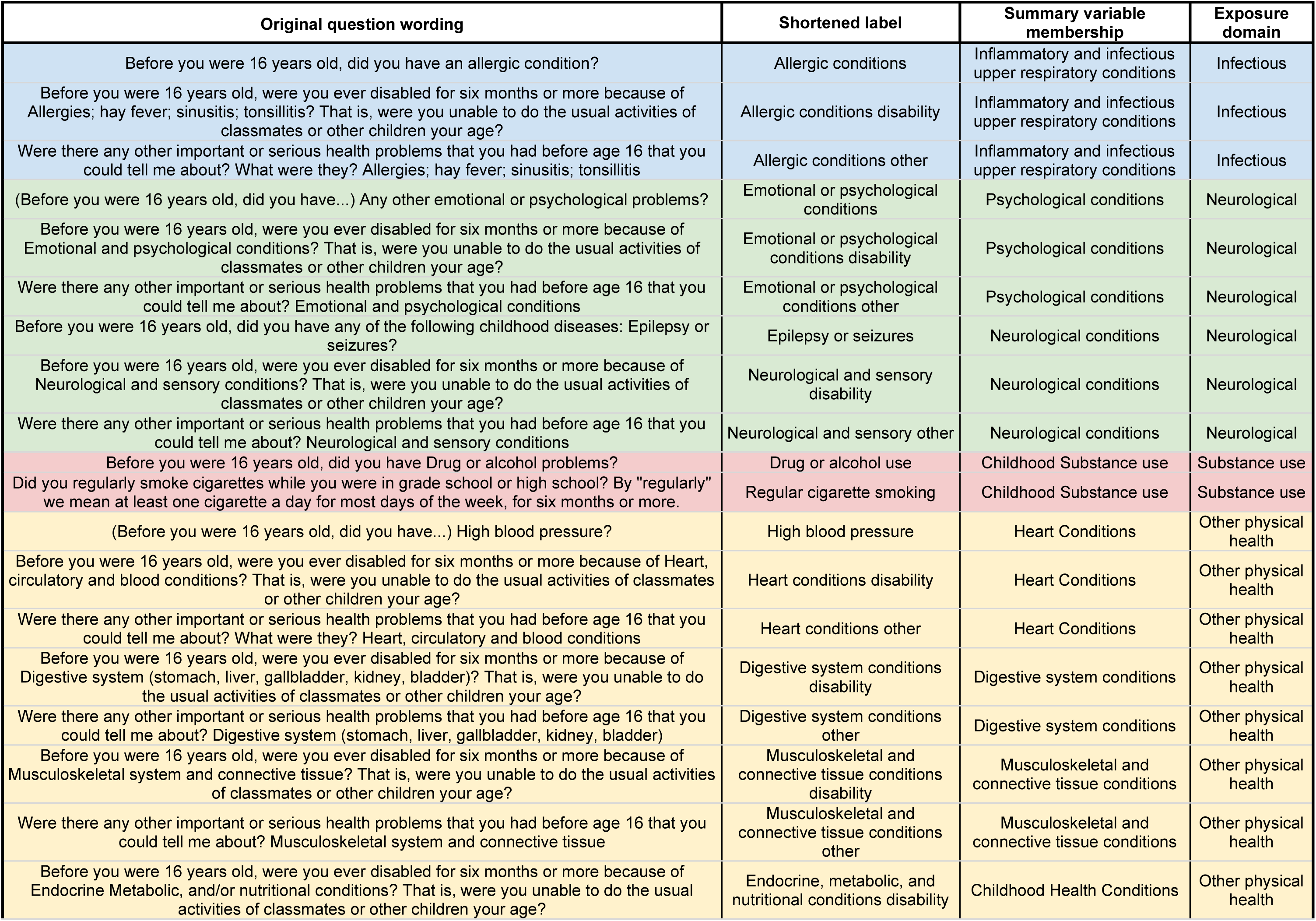
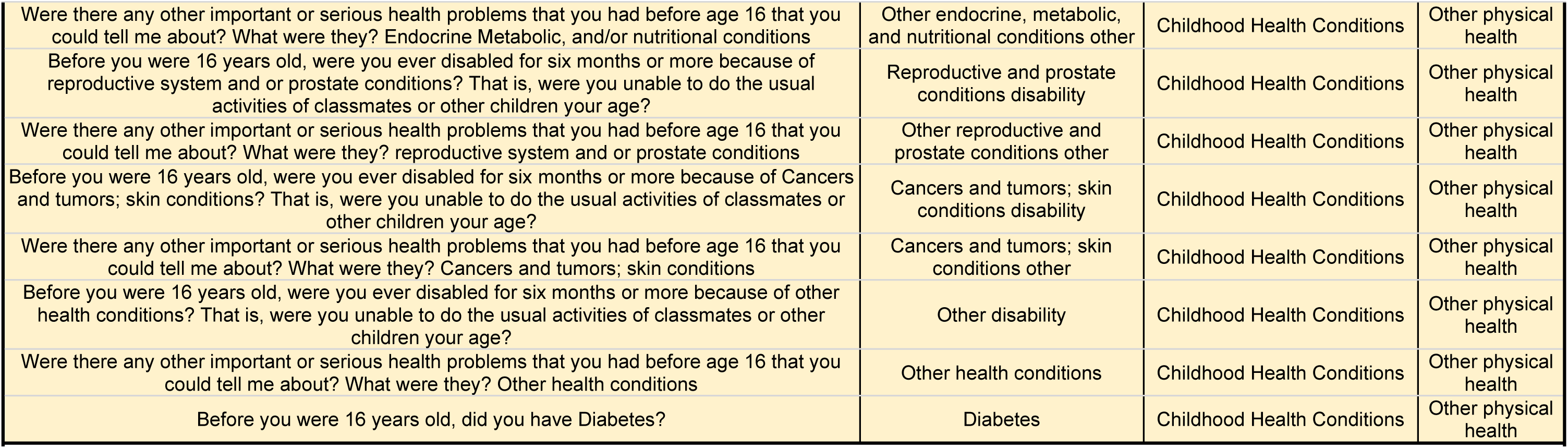
Table showing original question wording, shortened label for internal analysis, summary variable membership, and exposure domain for the eight summary exposure variables created in this analysis. Original question wording refers to the exact wording used in the retrospective questionnaire of exposures or conditions before age 16 in the 2008-2014 HRS waves. The shortened label is the original variable name used during internal analysis. The column "summary variable membership" displays the name of the new summary exposure variable created for this analysis. "Exposure domain" is the overarching category to which the newly generated summary variable belongs. Rows are color-coded based on exposure domain membership.

**Supplemental Table 2.** Characteristics of the Included vs. Excluded Study Individual s in Health and Retirement Study (HRS) sample 2008-2018.

| Characteristic | N | Overall<br>N = 22,018 | Excluded<br>N = 5,509 | Included<br>N = 16,509 | P-value<sup>2</sup> |
| --- | --- | --- | --- | --- | --- |
| **Cognition Status at baseline** | 22,018 |  |  |  | <0.001 |
| Normal Cognition |  | 17,698 (80%) | 1,189 (64%) | 16,509 (82%) |  |
| Cognitively impaired non-dementia | 3,473 (16%) | 3,473 (63%) | 0 (0%) |  |  |
| Dementia |  | 847 (3.8%) | 847 (15%) | 0 (0%) |  |
| **Cognition Status at follow-up** | 22,018 |  |  |  | <0.001 |
| Normal Cognition |  | 15,482 (70%) | 2,246 (41%) | 13,236 (80%) |  |
| Cognitively impaired non-dementia | 4,167 (19%) | 1,772 (32%) | 2,395 (15%) |  |  |
| Dementia |  | 2,369 (11%) | 1,491 (27%) | 878 (5.3%) |  |
| **Time from baseline to end of follow-up (years)** | 22,018 |  |  |  | <0.001 |
| 0 |  | 1,862 (8.5%) | 1,862 (100%) | 0 (0%) |  |
| 2 |  | 1,588 (7.2%) | 463 (8.4%) | 1,125 (6.8%) |  |
| 4 |  | 2,007 (9.1%) | 562 (10%) | 1,445 (8.8%) |  |
| 6 |  | 2,783 (13%) | 615 (11%) | 2,168 (13%) |  |
| 8 |  | 5,995 (27%) | 1,017 (18%) | 4,978 (30%) |  |
| 10 |  | 7,783 (35%) | 990 (18%) | 6,793 (41%) |  |
| **Age at baseline** | 22,018 | 64 (12) | 69 (13) | 63 (11) | <0.001 |
| **Proxy status at baseline** | 22,018 |  |  |  | <0.001 |
| No proxy |  | 21,684 (98%) | 5,328 (97%) | 16,356 (99%) |  |
| Proxy |  | 334 (1.5%) | 181 (3.3%) | 153 (0.9%) |  |
| **Proxy status at follow-up** | 22,018 |  |  |  | <0.001 |
| No proxy |  | 20,436 (93%) | 4,739 (86%) | 15,697 (95%) |  |
| Proxy |  | 1,582 (7.2%) | 770 (14%) | 812 (4.9%) |  |
| **Personal educational attainment (years)** | 22,018 | 12.8 (3.1) | 11.1 (3.7) | 13.3 (2.7) | <0.001 |
| **Sex** | 22,018 |  |  |  | <0.001 |
| Male |  | 9,133 (41%) | 2,396 (43%) | 6,737 (41%) |  |
| Female |  | 12,885 (59%) | 3,113 (57%) | 9,772 (59%) |  |
| **Race/ethnicity** | 22,018 |  |  |  | <0.001 |
| Non-Hispanic White |  | 14,581 (66%) | 2,964 (54%) | 11,617 (70%) |  |
| Non-Hispanic Black |  | 3,896 (18%) | 1,370 (25%) | 2,526 (15%) |  |
| Hispanic |  | 2,816 (13%) | 990 (18%) | 1,826 (11%) |  |
| Non-Hispanic Other |  | 725 (3.3%) | 185 (3.4%) | 540 (3.3%) |  |
| **Parental highest degree of education** | 22,018 |  |  |  | <0.001 |
| >= High School |  | 17,742 (81%) | 3,733 (68%) | 14,009 (85%) |  |
| < High School |  | 4,276 (19%) | 1,776 (32%) | 2,500 (15%) |  |
| **Infectious** |  |  |  |  |  |
| **Allergies; hay fever; sinusitis; tonsillitis** | 21,643 |  |  |  | <0.001 |
| No |  | 19,071 (88%) | 4,830 (91%) | 14,241 (87%) |  |
| Yes |  | 2,572 (12%) | 485 (9.1%) | 2,087 (13%) |  |
| **Measles before age 16** | 17,291 |  |  |  | <0.001 |
| No |  | 4,044 (23%) | 1,236 (26%) | 2,808 (22%) |  |
| Yes |  | 13,247 (77%) | 3,525 (74%) | 9,722 (78%) |  |
| **Mumps before age 16** | 17,222 |  |  |  | <0.001 |
| No |  | 6,934 (40%) | 2,028 (42%) | 4,906 (39%) |  |
| Yes |  | 10,288 (60%) | 2,757 (58%) | 7,531 (61%) |  |
| **Chickenpox before age 16** | 17,209 |  |  |  | <0.001 |
| No |  | 4,186 (24%) | 1,451 (31%) | 2,735 (22%) |  |
| Yes |  | 13,023 (76%) | 3,264 (69%) | 9,759 (78%) |  |
| **Neurological** |  |  |  |  |  |
| **Headaches or migraines before age 16** | 17,938 |  |  |  | 0.003 |
| No |  | 16,733 (93%) | 4,601 (92%) | 12,132 (94%) |  |
| Yes |  | 1,205 (6.7%) | 381 (7.6%) | 824 (6.4%) |  |
| **Ear problems before age 16** | 17,926 |  |  |  | 0.007 |
| No |  | 16,329 (91%) | 4,578 (92%) | 11,751 (91%) |  |
| Yes |  | 1,597 (8.9%) | 398 (8.0%) | 1,199 (9.3%) |  |
| **Depression before age 16** | 17,930 |  |  |  | 0.006 |
| No |  | 17,320 (97%) | 4,780 (96%) | 12,540 (97%) |  |
| Yes |  | 610 (3.4%) | 199 (4.0%) | 411 (3.2%) |  |
| **Emotional and psychological conditions** | 21,636 |  |  |  | 0.024 |
| No |  | 21,113 (98%) | 5,163 (97%) | 15,950 (98%) |  |
| Yes |  | 523 (2.4%) | 151 (2.8%) | 372 (2.3%) |  |
| **Neurological and sensory conditions** | 18,012 |  |  |  | 0.15 |
| No |  | 17,493 (97%) | 4,858 (97%) | 12,635 (97%) |  |
| Yes |  | 519 (2.9%) | 129 (2.6%) | 390 (3.0%) |  |
| **Childhood-learning problems** | 21,670 |  |  |  | <0.001 |
| No |  | 20,884 (96%) | 5,004 (94%) | 15,880 (97%) |  |
| Yes |  | 786 (3.6%) | 318 (6.0%) | 468 (2.9%) |  |
| **Concussion or severe head injury in childhood** | 21,641 |  |  |  | <0.001 |
| No |  | 19,699 (91%) | 4,921 (93%) | 14,778 (91%) |  |
| Yes |  | 1,942 (9.0%) | 395 (7.4%) | 1,547 (9.5%) |  |
| **Respiratory** |  |  |  |  |  |
| **Parents/guardians smoke** | 17,876 |  |  |  | <0.001 |
| No, none of them |  | 6,422 (36%) | 2,012 (41%) | 4,410 (34%) |  |
| Yes, one of them |  | 7,935 (44%) | 2,183 (44%) | 5,752 (45%) |  |
| Yes, both |  | 3,519 (20%) | 767 (15%) | 2,752 (21%) |  |
| **Asthma before age 16** | 17,933 |  |  |  | 0.6 |
| No |  | 16,985 (95%) | 4,708 (95%) | 12,277 (95%) |  |
| Yes |  | 948 (5.3%) | 271 (5.4%) | 677 (5.2%) |  |
| **Respiratory disorder before age 16** | 17,915 |  |  |  | <0.001 |
| No |  | 15,648 (87%) | 4,481 (90%) | 11,167 (86%) |  |
| Yes |  | 2,267 (13%) | 495 (9.9%) | 1,772 (14%) |  |
| Missing |  | 4,103 | 533 | 3,570 |  |
| **Substance use/Behavioral** |  |  |  |  |  |
| **Drugs, alcohol, or smoking use before age 16** | 18,686 |  |  |  | 0.016 |
| No |  | 14,468 (77%) | 3,983 (79%) | 10,485 (77%) |  |
| Yes |  | 4,218 (23%) | 1,082 (21%) | 3,136 (23%) |  |
| **Sensory** |  |  |  |  |  |
| **Difficulty seeing before age 16** | 17,938 |  |  |  | 0.028 |
| No |  | 16,887 (94%) | 4,657 (94%) | 12,230 (94%) |  |
| Yes |  | 1,051 (5.9%) | 323 (6.5%) | 728 (5.6%) |  |
| **Speech impairment before age 16** | 17,945 |  |  |  | 0.5 |
| No |  | 17,357 (97%) | 4,813 (97%) | 12,544 (97%) |  |
| Yes |  | 588 (3.3%) | 170 (3.4%) | 418 (3.2%) |  |
| **Other Physical Health** |  |  |  |  |  |
| **Heart, circulatory and blood conditions** | 17,978 |  |  |  | 0.4 |
| No |  | 17,238 (96%) | 4,776 (96%) | 12,462 (96%) |  |
| Yes |  | 740 (4.1%) | 193 (3.9%) | 547 (4.2%) |  |
| **Childhood health conditions** | 21,644 |  |  |  | <0.001 |
| No |  | 17,939 (83%) | 4,601 (87%) | 13,338 (82%) |  |
| Yes |  | 3,705 (17%) | 715 (13%) | 2,990 (18%) |  |
| **Childhood disability** | 21,663 |  |  |  | 0.2 |
| No |  | 20,826 (96%) | 5,098 (96%) | 15,728 (96%) |  |
| Yes |  | 837 (3.9%) | 221 (4.2%) | 616 (3.8%) |  |
| **Stomach problems before age 16** | 17,931 |  |  |  | 0.4 |
| No |  | 17,112 (95%) | 4,740 (95%) | 12,372 (96%) |  |
| Yes |  | 819 (4.6%) | 239 (4.8%) | 580 (4.5%) |  |
| **Rate health as child** | 17,879 |  |  |  | <0.001 |
| Excellent |  | 8,835 (49%) | 2,115 (43%) | 6,720 (52%) |  |
| Very Good |  | 5,014 (28%) | 1,412 (28%) | 3,602 (28%) |  |
| Good |  | 2,784 (16%) | 971 (20%) | 1,813 (14%) |  |
| Fair |  | 958 (5.4%) | 349 (7.0%) | 609 (4.7%) |  |
| Poor |  | 288 (1.6%) | 124 (2.5%) | 164 (1.3%) |  |
| **Digestive system (stomach, liver, gallbladder, kidney, bladder)** | 21,635 |  |  |  | <0.001 |
| No |  | 20,962 (97%) | 5,201 (98%) | 15,761 (97%) |  |
| Yes |  | 673 (3.1%) | 112 (2.1%) | 561 (3.4%) |  |
| **Musculoskeletal system and connective tissue** | 21,635 |  |  |  | 0.009 |
| No |  | 20,891 (97%) | 5,161 (97%) | 15,730 (96%) |  |
| Yes |  | 744 (3.4%) | 153 (2.9%) | 591 (3.6%) |  |
| **Child Home Environment** |  |  |  |  |  |
| **Lived in rural area as a child** | 21,177 |  |  |  | <0.001 |
| No |  | 11,750 (55%) | 2,428 (48%) | 9,322 (58%) |  |
| Yes |  | 9,427 (45%) | 2,617 (52%) | 6,810 (42%) |  |
| **Missed school as a child** | 17,891 |  |  |  | 0.019 |
| No |  | 15,938 (89%) | 4,465 (90%) | 11,473 (89%) |  |
| Yes |  | 1,953 (11%) | 496 (10.0%) | 1,457 (11%) |  |
| **Rate family financial situation** | 6,421 |  |  |  | <0.001 |
| Poor |  | 1,904 (30%) | 494 (36%) | 1,410 (28%) |  |
| About Avg |  | 3,856 (60%) | 745 (54%) | 3,111 (62%) |  |
| Pretty well off financially |  | 661 (10%) | 136 (9.9%) | 525 (10%) |  |
| **Live with grandparents during childhood** | 6,472 |  |  |  | 0.2 |
| No |  | 4,715 (73%) | 994 (72%) | 3,721 (73%) |  |
| Yes |  | 1,757 (27%) | 395 (28%) | 1,362 (27%) |  |
| **Father unemployed during childhood** | 6,381 |  |  |  | <0.001 |
| Never lived with Father/Father was not alive | 481 (7.5%) | 138 (10%) | 343 (6.8%) |  |  |
| No |  | 4,773 (75%) | 965 (71%) | 3,808 (76%) |  |
| Yes |  | 1,127 (18%) | 254 (19%) | 873 (17%) |  |
| **Mother work during childhood** | 7,172 |  |  |  | <0.001 |
| Not at all |  | 2,472 (34%) | 630 (37%) | 1,842 (34%) |  |
| All of the time |  | 1,991 (28%) | 517 (30%) | 1,474 (27%) |  |
| Some of the time |  | 2,709 (38%) | 558 (33%) | 2,151 (39%) |  |
| **Family get financial help in childhood** | 6,341 |  |  |  | 0.9 |
| No |  | 5,094 (80%) | 1,086 (80%) | 4,008 (80%) |  |
| Yes |  | 1,247 (20%) | 269 (20%) | 978 (20%) |  |

*CIND: Cognitive Impairment Non-Dementia; HRS: Health and Retirement Study; BP: Blood Pressure; SD: Standard Deviation;*
Notes: **[1]** Mean (SD) was calculated for continuous variables, Count(%) was calculated for categorical variables **[2]** Fisher's exact test was used to calculate p-values; Fisher's Exact Test for Count Data with simulated p-value (based on 2000 replicates). All P-values were interpreted as differences among groups. **[3]** Exposure sample sizes were allowed to vary by responses and missingness from the HRS survey. **[4]** Exposures were retrospectively analyzed from a self-reported questionnaire completed once within the 2008-2014 waves of the HRS. **[5]** Two additional years of HRS cognitive data on the same Individuals were included (2016 & 2018 HRS waves cognitive measures only). With the addition of the 2016 and 2018 cognition measures, our cognitive data spans from 2008 to 2018.

**Supplemental Table 3.** Crude Logistic Regression of Incident Cognitive Impairment Non-Dementia or Dementia vs. Normal Cognition and Exposures or Conditions before Age 16 in the Health and Retirement Study (HRS) 2008-2018.

|  |  |  | Dementia vs. Normal cognition |  |  |  | Cognitively impaired non-dementia vs. Normal cognition |  |  |  |
| --- | --- | --- | --- | --- | --- | --- | --- | --- | --- | --- |
|  | Exposed (N) | Responded (N) | OR | CI | P-value | FDR | OR | CI | P-value | FDR |
| Exposures before age 16 |  |  |  |  |  |  |  |  |  |  |
| Infectious |  |  |  |  |  |  |  |  |  |  |
| Measles (Yes) | 9,722 | 12,530 | 1.491 | (1.225, 1.829) | <0.001 | <0.001 | 1.199 | (1.066, 1.352) | 0.003 | 0.005 |
| Mumps (Yes) | 7,531 | 12,437 | 1.112 | (0.952, 1.302) | 0.182 | 0.252 | 1.002 | (0.908, 1.107) | 0.961 | 0.961 |
| Chicken pox (Yes) | 9,759 | 12,494 | 0.789 | (0.663, 0.943) | 0.008 | 0.015 | 0.768 | (0.687, 0.86) | <0.001 | <0.001 |
| Allergies; hay fever; sinusitis; tonsillitis (Yes) | 2,087 | 16,328 | 0.75 | (0.596, 0.934) | 0.012 | 0.021 | 0.704 | (0.609, 0.812) | <0.001 | <0.001 |
| Neurological |  |  |  |  |  |  |  |  |  |  |
| Headaches or migraines before age 16 (Yes) | 824 | 12,956 | 1.229 | (0.921, 1.612) | 0.148 | 0.218 | 0.834 | (0.675, 1.02) | 0.084 | 0.105 |
| Ear problems before age 16 (Yes) | 1,199 | 12,950 | 0.983 | (0.753, 1.264) | 0.899 | 0.926 | 0.972 | (0.823, 1.143) | 0.737 | 0.807 |
| Depression before age 16 (Yes) | 411 | 12,951 | 0.842 | (0.516, 1.296) | 0.463 | 0.56 | 1.036 | (0.788, 1.342) | 0.794 | 0.843 |
| Emotional and psychological conditions (Yes) | 372 | 16,322 | 0.959 | (0.58, 1.489) | 0.861 | 0.926 | 0.953 | (0.7, 1.272) | 0.752 | 0.811 |
| Neurological and sensory conditions (Yes) | 390 | 13,025 | 0.897 | (0.556, 1.368) | 0.633 | 0.705 | 0.751 | (0.547, 1.008) | 0.065 | 0.088 |
| Childhood learning problems (Yes) | 468 | 16,348 | 0.969 | (0.608, 1.462) | 0.886 | 0.926 | 1.668 | (1.325, 2.081) | <0.001 | <0.001 |
| Childhood concussion or severe head injury (Yes) | 1,547 | 16,325 | 0.849 | (0.658, 1.078) | 0.192 | 0.255 | 0.801 | (0.682, 0.937) | 0.006 | 0.01 |
| Respiratory |  |  |  |  |  |  |  |  |  |  |
| Parents or guardians smoke <sup>1</sup> | 8,504 | 12,914 |  |  |  |  |  |  |  |  |
| Yes, one of them | 5,752 |  | 0.943 | (0.802, 1.108) | 0.474 | 0.564 | 1.028 | (0.925, 1.143) | 0.607 | 0.687 |
| Yes, both | 2,752 |  | 0.494 | (0.389, 0.622) | <0.001 | <0.001 | 0.651 | (0.565, 0.748) | <0.001 | <0.001 |
| Asthma before age 16 (Yes) | 677 | 12,954 | 1.003 | (0.708, 1.383) | 0.984 | 0.984 | 0.991 | (0.797, 1.222) | 0.936 | 0.95 |
| Respiratory disorder before age 16 (Yes) | 1,772 | 12,939 | 0.849 | (0.674, 1.056) | 0.152 | 0.218 | 0.698 | (0.598, 0.81) | <0.001 | <0.001 |
| Behavioral/substance use |  |  |  |  |  |  |  |  |  |  |
| Drugs, alcohol, or smoking use before age 16 (Yes) | 3,136 | 13,621 | 0.79 | (0.655, 0.947) | 0.012 | 0.021 | 1.005 | (0.9, 1.121) | 0.926 | 0.95 |
| Sensory |  |  |  |  |  |  |  |  |  |  |
| Speech impairment before age 16 (Yes) | 418 | 12,962 | 0.877 | (0.55, 1.326) | 0.556 | 0.643 | 0.788 | (0.584, 1.042) | 0.106 | 0.13 |
| Difficulty seeing before age 16 (Yes) | 728 | 12,958 | 1.223 | (0.899, 1.629) | 0.182 | 0.252 | 0.856 | (0.686, 1.059) | 0.161 | 0.195 |
| Other Physical Health |  |  |  |  |  |  |  |  |  |  |
| Heart, circulatory and blood conditions (Yes) | 547 | 13,009 | 0.76 | (0.492, 1.12) | 0.188 | 0.255 | 0.796 | (0.614, 1.017) | 0.075 | 0.099 |
| Childhood health conditions (Yes) | 2,923 | 16,327 | 0.904 | (0.75, 1.083) | 0.281 | 0.36 | 0.813 | (0.72, 0.915) | 0.001 | 0.001 |
| Childhood Disability (Yes) | 616 | 16,344 | 0.954 | (0.642, 1.364) | 0.805 | 0.881 | 1.276 | (1.026, 1.572) | 0.025 | 0.038 |
| Stomach problems before age 16 (Yes) | 580 | 12,952 | 1.108 | (0.773, 1.54) | 0.559 | 0.643 | 0.896 | (0.702, 1.13) | 0.364 | 0.419 |
| Rate health as child <sup>2</sup> | 6,188 | 12,908 |  |  |  |  |  |  |  |  |
| Very good | 3,602 |  | 1.485 | (1.244, 1.771) | <0.001 | <0.001 | 1.133 | (1.013, 1.266) | 0.029 | 0.042 |
| Good | 1,813 |  | 1.86 | (1.506, 2.287) | <0.001 | <0.001 | 1.363 | (1.187, 1.562) | <0.001 | <0.001 |
| Fair | 609 |  | 1.779 | (1.271, 2.436) | 0.001 | 0.001 | 1.224 | (0.975, 1.524) | 0.076 | 0.099 |
| Poor | 164 |  | 3.298 | (1.989, 5.218) | <0.001 | <0.001 | 1.434 | (0.938, 2.127) | 0.083 | 0.105 |
| Digestive system (stomach, liver, gallbladder, kidney, bladder) (Yes) | 561 | 16,322 | 1.015 | (0.687, 1.445) | 0.936 | 0.95 | 0.78 | (0.595, 1.005) | 0.063 | 0.086 |
| Musculoskeletal system and connective tissue (Yes) | 591 | 16,321 | 0.598 | (0.368, 0.914) | 0.026 | 0.042 | 0.765 | (0.589, 0.979) | 0.039 | 0.055 |
| Child Home Environment |  |  |  |  |  |  |  |  |  |  |
| Lived rural area during school (Yes) | 6,810 | 16,132 | 1.154 | (1.001, 1.329) | 0.047 | 0.075 | 1.262 | (1.155, 1.379) | <0.001 | <0.001 |
| Childhood- Missed school (Yes) | 1,457 | 12,930 | 1.248 | (0.997, 1.547) | 0.048 | 0.075 | 1.033 | (0.888, 1.198) | 0.666 | 0.741 |
| Live with grandparents during childhood | 1,362 | 5,083 | 0.673 | (0.33, 1.257) | 0.241 | 0.314 | 1.125 | (0.912, 1.382) | 0.266 | 0.316 |
| Father unemployed during childhood <sup>3</sup> | 1,216 | 5,024 |  |  |  |  |  |  |  |  |
| Never lived with father/father not alive | 343 |  | 0.55 | (0.089, 1.793) | 0.409 | 0.504 | 1.806 | (1.298, 2.469) | <0.001 | 0.001 |
| Yes | 873 |  | 1.047 | (0.495, 2.007) | 0.898 | 0.926 | 1.362 | (1.069, 1.722) | 0.011 | 0.017 |
| Mother work during childhood <sup>4</sup> | 3,316 | 5,467 |  |  |  |  |  |  |  |  |
| Not at all | 1,842 |  | 1.257 | (0.777, 2.038) | 0.351 | 0.441 | 1.114 | (0.903, 1.373) | 0.312 | 0.365 |
| All of the time | 1,474 |  | 0.63 | (0.326, 1.158) | 0.151 | 0.218 | 1.257 | (1.012, 1.559) | 0.038 | 0.055 |
| Family get financial help in childhood (Yes) | 978 | 4,986 | 0.825 | (0.377, 1.612) | 0.601 | 0.679 | 1.355 | (1.079, 1.689) | 0.008 | 0.012 |
| Rate family financial situation- SES <sup>5</sup> | 1,935 | 5,046 |  |  |  |  |  |  |  |  |
| Poor | 1,410 |  | 2.414 | (1.353, 4.315) | 0.003 | 0.005 | 1.444 | (1.174, 1.77) | <0.001 | 0.001 |
| Pretty well off financially | 525 |  | 2.071 | (0.864, 4.471) | 0.078 | 0.12 | 0.962 | (0.679, 1.333) | 0.823 | 0.861 |

HRS: Health and Retirement Study; CIND: Cognitive impairment Non-Dementia; OR: Odds Ratio; BP: Blood Pressure; SES: Socioeconomic status; <sup>1</sup>Reference group in parents or guardians smoke is "No, None of them"; <sup>2</sup>Reference group in Rate health as a child is "Excellent"; <sup>3</sup>Reference group in Father unemployed during childhood is "No"; <sup>4</sup>Reference group in Mother work during childhood is "Some of the time"; <sup>5</sup>Reference group in Rate family financial situation is "About average".
[1] Cognitive status was defined as the cognition status of the Individual at their most recent follow-up cognitive assessment visit (Cognitive impairment non-dementia, Dementia, or normal) [2] Exposures were retrospectively analyzed from a self-reported questionnaire completed once in the 2008-2014 waves of the HRS. [3] Two additional years of HRS cognitive data on the same Individual s were included (2016 & 2018 HRS waves cognitive measures only). With the addition of the 2016 and 2018 cognition measures, our cognitive data spans from 2008 to 2018. [4] "Responded" is the total number of people with a valid measurement record for that exposure. "Exposed" is the count of people who responded yes to that exposure. [5] The P-value was calculated using logistic regression and interpreted as differences between groups.

**Supplemental Table 4.** Logistic regression of Incident Cognitive Impairment (any) and Exposures or Conditions before Age 16 in the Health and Retirement Study (HRS) 2008-2018.

|  | Exposed (N) | Responded (N) | Any Cognitive impairment vs. Normal Cognition |  |  |  |
| --- | --- | --- | --- | --- | --- | --- |
|  |  |  | OR | (95% CI) | P-value | FDR |
| Exposures before age 16 |  |  |  |  |  |  |
| Infectious |  |  |  |  |  |  |
| Measles (Yes) | 9,573 | 12,311 | 0.845 | (0.747, 0.957) | 0.008 | 0.05143 |
| Mumps (Yes) | 7,420 | 12,221 | 0.893 | (0.809, 0.985) | 0.024 | 0.09 |
| Chicken pox (Yes) | 9,604 | 12,275 | 0.912 | (0.813, 1.023) | 0.113 | 0.22109 |
| Allergies; hay fever; sinusitis; tonsillitis (Yes) | 2,048 | 12,939 | 0.984 | (0.857, 1.128) | 0.818 | 0.87643 |
| Overall odds ratio |  |  | 0.902 | (0.851, 0.956) | 0.001 | 0.015 |
| Neurological |  |  |  |  |  |  |
| Headaches or migraines before age 16 (Yes) | 813 | 12,715 | 1.135 | (0.934, 1.373) | 0.196 | 0.3528 |
| Ear problems before age 16 (Yes) | 1,186 | 12,708 | 1.086 | (0.923, 1.276) | 0.316 | 0.43364 |
| Depression before age 16 (Yes) | 398 | 12,704 | 1.742 | (1.33, 2.26) | 0.000 | 0 |
| Emotional and psychological conditions (Yes) | 363 | 12,725 | 1.343 | (0.999, 1.783) | 0.046 | 0.12177 |
| Neurological and sensory conditions (Yes) | 384 | 16,254 | 1.106 | (0.827, 1.461) | 0.487 | 0.57671 |
| Childhood learning problems (Yes) | 463 | 16,101 | 2.125 | (1.675, 2.681) | 0.000 | 0.000 |
| Childhood concussion or severe head injury (Yes) | 1,530 | 16,083 | 1.152 | (0.987, 1.34) | 0.071 | 0.16816 |
| Overall odds ratio |  |  | 1.331 | (1.096, 1.615) | 0.004 | 0.03 |
| Respiratory |  |  |  |  |  |  |
| Parents or guardians smoke <sup>1</sup> |  |  |  |  |  |  |
| Yes, one of them | 5,654 | 12,694 | 1.002 | (0.903, 1.112) | 0.968 | 0.968 |
| Yes, both | 2,709 | 12,694 | 0.983 | (0.855, 1.129) | 0.811 | 0.87643 |
| Asthma before age 16 (Yes) | 663 | 12,710 | 1.36 | (1.101, 1.67) | 0.004 | 0.03 |
| Respiratory disorder before age 16 (Yes) | 1,741 | 12,701 | 1.055 | (0.913, 1.216) | 0.467 | 0.57671 |
| Overall odds ratio |  |  | 1.067 | (0.947, 1.203) | 0.284 | 0.43364 |
| Behavioral/substance use |  |  |  |  |  |  |
| Drugs, alcohol, or smoking use before age 16 (Yes) | 3,085 | 16,254 | 1.134 | (1.014, 1.267) | 0.026 | 0.09 |
| Sensory |  |  |  |  |  |  |
| Speech impairment before age 16 (Yes) | 410 | 12,713 | 1.243 | (0.939, 1.627) | 0.121 | 0.22688 |
| Difficulty seeing before age 16 (Yes) | 716 | 12,714 | 1.035 | (0.844, 1.264) | 0.735 | 0.82688 |
| Overall odds ratio |  |  | 1.107 | (0.932, 1.315) | 0.248 | 0.39857 |
| Other Physical Health |  |  |  |  |  |  |
| Heart, circulatory and blood conditions (Yes) | 537 | 12,788 | 0.881 | (0.69, 1.115) | 0.3 | 0.43364 |
| Childhood health conditions (Yes) |  |  | 0.881 | (0.785, 0.988) | 0.031 | 0.099 |
| Childhood Disability (Yes) | 611 | 16,095 | 1.231 | (0.993, 1.518) | 0.054 | 0.135 |
| Stomach problems before age 16 (Yes) | 567 | 12,714 | 1.212 | (0.963, 1.516) | 0.096 | 0.216 |
| Rate health as child <sup>2</sup> |  |  |  |  |  |  |
| Very good | 3,556 | 12,717 | 1.025 | (0.919, 1.143) | 0.652 | 0.75231 |
| Good | 1,784 | 12,717 | 1.177 | (1.028, 1.347) | 0.018 | 0.09 |
| Fair | 591 | 12,717 | 1.25 | (1.004, 1.548) | 0.044 | 0.12177 |
| Poor | 161 | 12,717 | 1.754 | (1.196, 2.544) | 0.003 | 0.03 |
| Digestive system (stomach, liver, gallbladder, kidney, bladder) (Yes) | 556 | 16,254 | 0.894 | (0.699, 1.133) | 0.363 | 0.48044 |
| Musculoskeletal system and connective tissue (Yes) | 588 | 16,254 | 0.814 | (0.631, 1.038) | 0.104 | 0.21886 |
| **Overall odds ratio** |  |  | 1.063 | (0.943, 1.199) | 0.318 | 0.43364 |
| **Child Home Environment** |  |  |  |  |  |  |
| Lived rural area during school (Yes) | 9,185 | 15,884 | 1.107 | (1.014, 1.209) | 0.024 | 0.09 |
| Childhood- Missed school (Yes) | 1,431 | 12,688 | 1.076 | (0.931, 1.24) | 0.318 | 0.43364 |
| Live with grandparents during childhood | 1,275 | 4,854 | 0.925 | (0.748, 1.139) | 0.467 | 0.57671 |
| Father unemployed during childhood <sup>3</sup> |  |  |  |  |  |  |
| Never lived with father/father not alive | 323 | 4,804 | 1.128 | (0.802, 1.562) | 0.477 | 0.57671 |
| Yes | 841 | 4,804 | 1.216 | (0.955, 1.539) | 0.107 | 0.21886 |
| Mother work during childhood <sup>4</sup> |  |  |  |  |  |  |
| Not at all | 1,782 | 5,308 | 0.874 | (0.704, 1.082) | 0.217 | 0.37558 |
| All of the time | 1,435 | 5,308 | 0.993 | (0.799, 1.232) | 0.947 | 0.968 |
| Family get financial help in childhood (Yes) | 940 | 4,767 | 1.279 | (1.017, 1.599) | 0.033 | 0.099 |
| Rate family financial situation-SES <sup>5</sup> |  |  |  |  |  |  |
| Poor | 1,365 | 4,856 | 1.02 | (0.822, 1.262) | 0.857 | 0.89686 |
| Pretty well off financially | 501 | 4,856 | 1.222 | (0.875, 1.679) | 0.226 | 0.37667 |
| **Overall odds ratio** |  |  | 1.075 | (1.014, 1.139) | 0.015 | 0.08438 |

HRS: Health and Retirement Study; CIND: Cognitive impairment Non-Dementia; OR: Odds Ratio; BP: Blood Pressure; SES: Socioeconomic status; <sup>1</sup>Reference group in parents or guardians smoke is "No, None of them"; <sup>2</sup>Reference group in Rate health as child is "Excellent"; <sup>3</sup>Reference group in Father unemployed during childhood is "No"; <sup>4</sup>Reference group in Mother work during childhood is "Some of the time"; <sup>5</sup>Reference group in Rate family financial situation is "About average".

